# Prevalence and factors associated with tobacco and nicotine product use among adolescents in the Democratic Republic of the Congo: evidence from a cross-sectional national household survey

**DOI:** 10.64898/2026.05.01.26352215

**Authors:** Christelle Tchoupé, Didier Munguakonkwa Mirindi, Josiane Djiofack Tsague, Bruce Wife Nana Nana, Retselisitsoe Pokothoane, Grace Kyule, Samuel Iddi, Lyagamula Kisia, Olatunbosun Abolarin, Thompson Ademola, Akinsewa Akiode, Terefe Gelibo Agerfa, Emmanuel Kandate, David Kayembe, Patrice Milambo, Jean Chris Mampuya, Nelson Mbaya, Shukri F. Mohamed, Célestin Banza Lubaba Nkulu, Uche Okezie, Boscow Okumu, Ndelo Phanzu, Patrick Bakengela Shamba, Noreen Dadirai Mdege

## Abstract

**Background:** The initiation of tobacco and nicotine product use often occurs in adolescents. This necessitates monitoring of this behaviour in this population, particularly in countries such as the Democratic Republic of the Congo (DRC) where approximately 58% of the population is under 19 years of age. Our study assessed the prevalence of, and factors associated with use in the DRC.

**Methods:** We conducted a nationally representative, cross-sectional, household survey between March and May 2024 among adolescents aged 10 to 17 years. We estimated the prevalence of use of smoked and smokeless tobacco products, heated tobacco products, and nicotine products (i.e., electronic cigarettes and nicotine pouches). We used logistic regression to identify factors associated with current use of any tobacco product, smoked tobacco, and smokeless tobacco using adjusted odd ratios. All analyses included 95% confidence intervals (CI).

**Results:** Of the 4,675 adolescents who completed the survey, the prevalence of current use of any tobacco or nicotine product was 11.87% (95% CI: 6.93-19.58). This was 7.98% (95% CI: 4.23-14.55) for smoked tobacco products, 5.86% (95% CI: 3.42-9.87) for smokeless tobacco products, 0.11% (95% CI: 0.11-0.11) for heated tobacco products and 0.60% (95% CI: 0.10-3.40) for nicotine products. Boys were more likely to use tobacco than girls. Being enrolled in school and having both parents alive were protective from tobacco use. Having a male household head, a household head education level of at least secondary school, and exposure to tobacco smoking in public places increased the odds of being a tobacco user.

**Conclusions:** The DRC should strengthen policies that make tobacco and nicotine products less accessible or appealing to adolescents. This includes raising excise taxes; banning the sale of single cigarette sticks, small packets and flavoured products; and comprehensive smoke-free laws. Policies should account for factors that make adolescents more vulnerable product use.

**Key messages:** *What is already known on the topic:*

- The last survey on tobacco use among adolescents in the DRC was a school-based survey among 13-15-year-olds conducted in 2008, and only covered Kinshasa and Lubumbashi.

*What this study adds:*

- This survey provided national-level estimates that cover adolescents aged 10-17years, includes out-of-school adolescents, and covers both tobacco and nicotine products.
- It also identifies individual-, household-, and environmental-level factors that are associated with tobacco and nicotine product use among adolescents in the DRC.

*How this study might affect research, practice or policy:*

- By providing current and more comprehensive data, our study enhances policymakers’ ability to design evidence-based tobacco control interventions that are aimed at preventing the initiation and use of tobacco and nicotine products among adolescents in the DRC and other similar settings.

## Introduction

Tobacco use remains a critical global public health threat, contributing to more than 8 million deaths each year, including approximately 1.2 million non-smokers exposed to second-hand smoke [1]. Among adolescents aged 13–15 years, around 12% report using some form of tobacco, with prevalence estimates in low- and middle-income countries (LMICs) ranging from 11% to 13% [2]. In the Democratic Republic of Congo (DRC) approximately 58% of the population is under 19 years of age [3]. Such a high proportion of children and adolescents presents a strategic target for the tobacco industry, which employs tactics such as celebrity endorsements, social media influencers, advertising near schools, flavored products, and free samples to encourage uptake [4, 5]. These strategies promote the use of both traditional tobacco products and emerging nicotine products such as electronic cigarettes [6, 7]. Even some tobacco products that, traditionally, were not widely used by adolescents, such as shisha, are becoming increasingly popular among this population in Africa: many adolescents perceive them as safer than cigarettes [8].

The DRC ratified the WHO Framework Convention on Tobacco Control (WHO FCTC) in 2005, and adopted Law No. 18/035 in 2018, introducing key measures such as bans on smoking in public spaces, advertising and sponsorship restrictions, written health warnings on packaging, and prohibitions on sales to/and by minors [9, 10]. However, these legal provisions are currently poorly implemented, and regulatory gaps persist including lack of bans/restrictions on the sale of single cigarette sticks or small packets, flavored products, shisha, and emerging nicotine products such as electronic cigarettes. Point of sale product displays, and the sale of tobacco products in or near locations that are frequented by children and adolescents such as school/other educational facilities and playgrounds, are not prohibited.

The DRC has a significant gap in the monitoring of tobacco and nicotine product use. The currently available data on tobacco use among adolescents is from the 2008 Global Youth Tobacco Survey which was restricted to two urban areas, i.e., Kinshasa and Lubumbashi. In addition, this survey was conducted in schools and targeted those aged 13-15 years, despite the fact that in the DRC, 20% to 30% of children aged 6-17 are out of school [11], and the young people most likely to start smoking are those who are not actively enrolled in school [12]. Moreover, data from Sub-Saharan Africa indicate that the age at which adolescents begin smoking ranges from 7 to 16 years old [11, 13]. There is also no data on the use of emerging tobacco and nicotine products such as heated tobacco products or electronic cigarettes among adolescents in the DRC. To address this critical data gap, we conducted a nationally representative, cross-sectional, household survey to assess the prevalence of, and identify individual-, household-, and environmental-level factors that are associated with tobacco and nicotine product use among adolescents aged 10 to 17 years in the DRC. These data will enhance policymakers’ ability to design evidence-based tobacco control interventions that are aimed at preventing the initiation and use of tobacco and nicotine products in this population.

## Methods

### Study design and setting

This study was a nationally representative cross-sectional survey among adolescents aged 10 to 17 years. Data were collected from 144 enumeration areas selected using multistage, stratified cluster sampling. The sampling frame was drawn from the National Health Information System and is based on the Ministry of Public Health, Hygiene, and Prevention’s health pyramid. The 26 current provinces of the DRC were grouped into six strata aligned with the former six provinces of 1947–1963, i.e., Katanga, Kasaï, Léopoldville, Équateur, Orientale, and Kivu. Sixteen participating provinces were then randomly selected from the six strata [14]. Each selected province received an equal share of the household quota within its stratum.

In each of the selected provinces, three health zones (HZs), two rural and one urban, were randomly selected resulting in a total of 48 HZs. From each HZ, we randomly selected three health areas (HAs), two rural and one urban, yielding a total of 144 HAs. One village (in rural HAs) or avenue (in urban HAs), which were the enumeration areas (EAs), was randomly selected per HA. We then conducted household listing to identify households with adolescents aged 10–17 years in each participating EA and drew a random sample of households from those that were eligible. One adolescent aged 10-17 years was interviewed in each participating household, with random selection applied for households with more than one eligible adolescent.

### Study population

Adolescents aged 10–17 years old were eligible to participate in the study if they were a member of a participating household whose household head had provided household-level consent and consent for the participation of the selected adolescent. Adolescent assent (or consent for emancipated minors) was obtained prior to survey completion. Those unable to provide assent, or those with physical disabilities preventing oral survey participation were excluded.

### Sample size

The sample size was calculated using the United Nations formula [15], incorporating a 95% confidence level, a design effect of 1.5, and a 10% non-response adjustment, in line with previous studies in the DRC [16, 17]. An average household size was set to 5.25 [3], and an estimated tobacco use prevalence of 25%, informed by earlier findings from Kinshasa (22.3%) and Lubumbashi (24.6%) [11]. Key assumptions included an adolescent population share of 23%. Applying these parameters, the required minimum sample was 4,323. Full details of the calculation are provided in the study protocol [14].

### Survey materials

The survey comprised two questionnaires: a household questionnaire and an adolescent questionnaire. The development of the questionnaires has been described in detail in the study protocol [14]. The household questionnaire administered to the head of household comprised two modules: demographics and socio-economic status. The first captured information on household members, including sex, age, income, disability, marital status, education, and health insurance. The second assessed living conditions, including water sources, sanitation, cooking facilities, housing materials, and asset ownership.

The adolescent questionnaire comprised 12 modules designed to assess tobacco and nicotine product use, and individual, household, and environment-level factors associated with use. One module focused on socio-demographic characteristics, including age, sex, school status and grade level, weekly spending money, marital status, functional difficulties (e.g., vision, mobility, cognition), and tobacco use by family members and close friends. There were eight modules assessing tobacco and nicotine product use, covering smoked tobacco (i.e., manufactured cigarettes, roll-your-own cigarettes, shisha, and other smoked products such as cigars), smokeless tobacco (e.g., chewing tobacco, snuff), heated tobacco products, e-cigarettes, and nicotine pouches. For each product, data were collected on use patterns, frequency, quantity, dependence, age of initiation, context of use, and access. The questionnaire also included one module exploring knowledge, attitudes and perceptions related to tobacco products and their use, as well as exposure to pro- and anti-tobacco messaging. This module was administered only to adolescents who had ever used a tobacco or nicotine product. An additional module captured information on intentions to quit tobacco among users, and the last one was on second-hand smoke exposure, capturing information on exposure to tobacco smoke in homes and public spaces.

The survey instruments were developed in English. They were translated to French by a professional native French speaker, followed by independent back-translation by another native speaker unfamiliar with the original content. Considering the multiple local languages spoken in DRC, data collectors were trained to translate the questionnaires orally during fieldwork, enhancing accessibility and comprehension of study participants across provinces. The survey instruments and data collection processes were field tested prior to deployment and revised to address any issues identified such as ambiguous wording, technical challenges with electronic tools, and difficulties obtaining consent.

### Data collection

Data was collected between March and May 2024 by 80 fieldworkers (16 teams of 1 supervisor and 4 enumerators) who were deployed across the participating provinces, conducting surveys simultaneously. The questionnaires were programmed in SurveyCTO [18] on tablets, and interviewer-administered face-to-face. The secure electronic platform supported offline data entry and seamless online uploading to central servers. Interviews were conducted in the respondent’s preferred language (French, Lingala, Swahili, Tshiluba, or Kikongo) and lasted between 20 and 45 minutes. Interviewers worked in their home or resident provinces to minimize linguistic and cultural barriers.

Prior to data collection, all field staff completed a four-day training, including one day of field testing. Training covered study objectives, ethical consideration, survey tools, and mock interviews, with a written assessment at the end. Supervisors received additional instruction on logistics and quality control. Due to the country’s size, training was delivered in two phases: supervisors were trained in Kinshasa (4–7 March 2024), and enumerators received a standardized training in each selected province (20–23 March 2024). Fieldworker safety was ensured through check-ins, emergency protocols, and coordination with local authorities.

### Statistical analysis

All analyses were conducted using STATA version 18. Survey-weighted descriptive statistics were used to summarize household and adolescent characteristics and to estimate the prevalence of tobacco and nicotine product use. Frequencies and percentages were reported for categorical variables, while means/median, and standard deviations were calculated for continuous variables. Standard errors were provided alongside all point estimates. Prevalence estimates for ever and current use of each tobacco and nicotine product were calculated overall and disaggregated by age, sex, school attendance, residence type, household wealth quintile, marital status, and work engagement. "Ever use" was defined as having tried a product at least once in a lifetime, while "current use" referred to use within the past 30 days. Refusals and “don’t know” responses were treated as missing.

Survey-weighted logistic regression models were fitted to estimate adjusted odds ratios (aORs) with 95% confidence intervals for current use of any tobacco product, smoked tobacco, and smokeless tobacco. Due to small number of users, regression analyses were not conducted for current use of nicotine products (i.e., electronic cigarettes and nicotine pouches) or heated tobacco products. Covariates were structured according to the socio-ecological framework [19, 20], encompassing individual-level (e.g., sex, age, school attendance, employment status, parental survival), household-level (e.g., family smoking, household head’s gender and education), and environmental-level factors (e.g., teacher tobacco use, second-hand smoke exposure, rural/urban residence). Variable selection was informed by theoretical relevance, empirical evidence, and contribution to model performance. Multicollinearity was assessed using Variance Inflation Factors (VIF); variables with VIF >5 was reviewed, and redundant predictors were excluded.

All analyses accounted for sampling weights, clustering and stratification. Sampling weights were computed as the inverse of selection probabilities across all sampling stages and adjusted for non-response. We applied a calibration factor to align the weighted sample with target population distributions by strata.

### Participant and public involvement

Key stakeholders, including youth and adolescent advocates, policymakers, public health practitioners, tobacco control advocates, and academics were involved in the design, conduct, and dissemination of the study. This was achieved through one-on-one discussions, and co-design and validation workshops that informed study priorities, questionnaire development, and study implementation procedures, and validated the findings. More details of the activities are provided in the study protocol [14].

## Results

### Household and adolescent participation

Of the 15,864 households listed during the enumeration phase, 9,226 were eligible for inclusion. From these, 4,892 households were randomly selected for participation and 4,867 consented to participate (Figure 1). The household characteristics are provided in Supplementary Material Table 1. The consenting households had a total of 22,291 household members and 9,186 eligible adolescents. One adolescent aged 10-17 years was randomly selected to participate from each of the consenting households and 4,851 provided informed assent. Of these, 4,675 adolescents completed the survey and were included in the final analysis.

**Figure 1:**
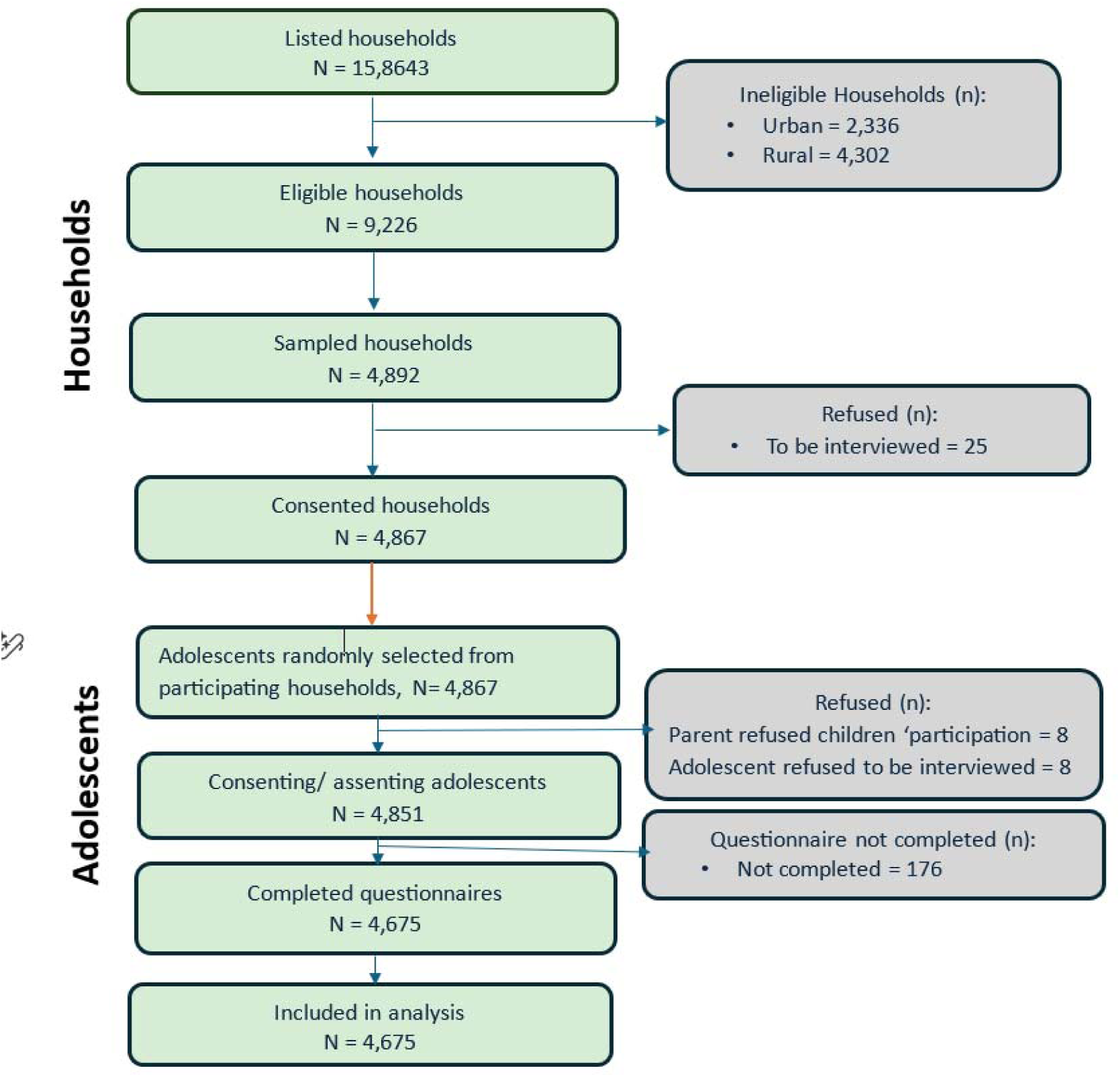
Flow of households and adolescents in the study.

### Adolescent characteristics

The mean age of participating adolescents was 13.0 years (Table 1). The majority (88.5%) were attending school, and the median weekly pocket money was approximately 3,075.5 CDF (boys: 3,348.0; girls: 2,751.4). Most adolescents (84.1%) did not engage in work, with 2.5% employed and 8.0% self-employed. Among those aged 15 years and older who reported their marital status, nearly all (97.1%) stated they were not in a union, while 1.8% were currently in a union. Christianity was the predominant religion (87.9%), and nearly all adolescents (96.4%) reported no functional disability. The majority reported their biological mother (87.7%) and father (82.1%) as alive. Most adolescents resided in rural areas (62.6%), and the largest regional representation was from Leopoldville (23.3%), followed by Kasai (19.6%) and Kivu (18.0%). Overall, the distribution of characteristics was similar between boys and girls.

**Table 1:**
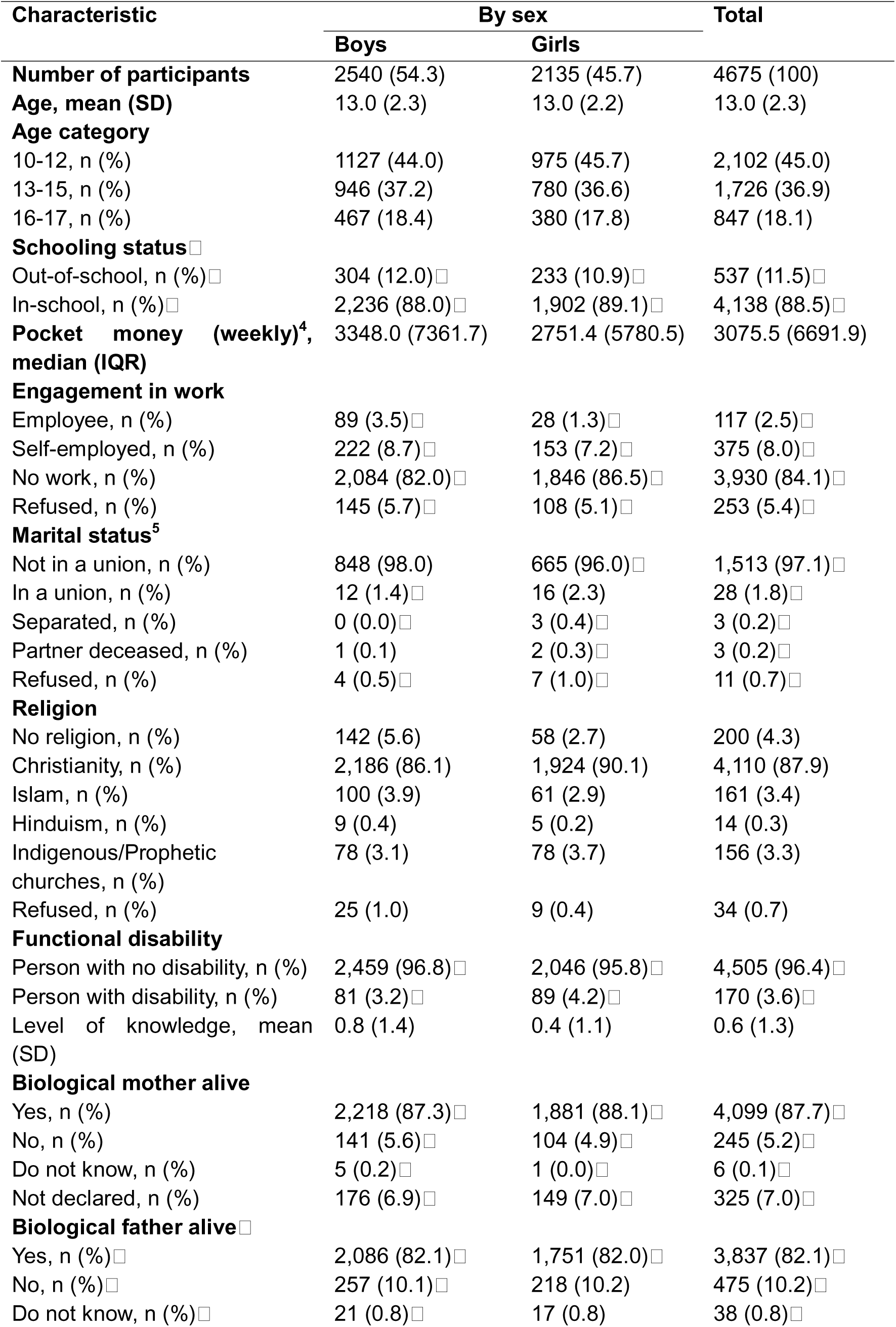
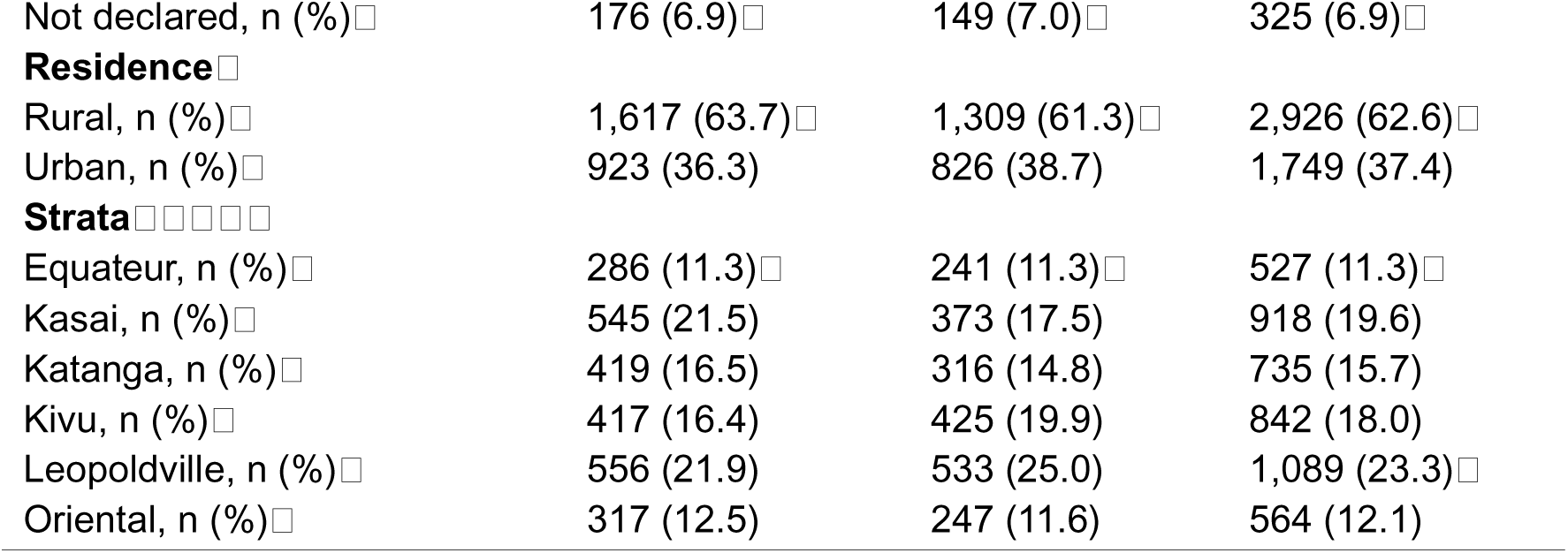
Adolescents characteristics.

| Characteristic | By sex |  | Total |
| --- | --- | --- | --- |
|  | Boys | Girls |  |
| **Number of participants** | 2540 (54.3) | 2135 (45.7) | 4675 (100) |
| **Age, mean (SD)** | 13.0 (2.3) | 13.0 (2.2) | 13.0 (2.3) |
| **Age category** |  |  |  |
| 10-12, n (%) | 1127 (44.0) | 975 (45.7) | 2,102 (45.0) |
| 13-15, n (%) | 946 (37.2) | 780 (36.6) | 1,726 (36.9) |
| 16-17, n (%) | 467 (18.4) | 380 (17.8) | 847 (18.1) |
| **Schooling status** □ |  |  |  |
| Out-of-school, n (%) □ | 304 (12.0) □ | 233 (10.9) □ | 537 (11.5) □ |
| In-school, n (%) □ | 2,236 (88.0) □ | 1,902 (89.1) □ | 4,138 (88.5) □ |
| **Pocket money (weekly)<sup>4</sup>, median (IQR)** | 3348.0 (7361.7) | 2751.4 (5780.5) | 3075.5 (6691.9) |
| **Engagement in work** |  |  |  |
| Employee, n (%) | 89 (3.5) □ | 28 (1.3) □ | 117 (2.5) □ |
| Self-employed, n (%) | 222 (8.7) □ | 153 (7.2) □ | 375 (8.0) □ |
| No work, n (%) | 2,084 (82.0) □ | 1,846 (86.5) □ | 3,930 (84.1) □ |
| Refused, n (%) | 145 (5.7) □ | 108 (5.1) □ | 253 (5.4) □ |
| **Marital status<sup>5</sup>** |  |  |  |
| Not in a union, n (%) | 848 (98.0) | 665 (96.0) □ | 1,513 (97.1) □ |
| In a union, n (%) | 12 (1.4) □ | 16 (2.3) | 28 (1.8) □ |
| Separated, n (%) | 0 (0.0) □ | 3 (0.4) □ | 3 (0.2) □ |
| Partner deceased, n (%) | 1 (0.1) | 2 (0.3) □ | 3 (0.2) □ |
| Refused, n (%) | 4 (0.5) □ | 7 (1.0) □ | 11 (0.7) □ |
| **Religion** |  |  |  |
| No religion, n (%) | 142 (5.6) | 58 (2.7) | 200 (4.3) |
| Christianity, n (%) | 2,186 (86.1) | 1,924 (90.1) | 4,110 (87.9) |
| Islam, n (%) | 100 (3.9) | 61 (2.9) | 161 (3.4) |
| Hinduism, n (%) | 9 (0.4) | 5 (0.2) | 14 (0.3) |
| Indigenous/Prophetic churches, n (%) | 78 (3.1) | 78 (3.7) | 156 (3.3) |
| Refused, n (%) | 25 (1.0) | 9 (0.4) | 34 (0.7) |
| **Functional disability** |  |  |  |
| Person with no disability, n (%) | 2,459 (96.8) □ | 2,046 (95.8) □ | 4,505 (96.4) □ |
| Person with disability, n (%) | 81 (3.2) □ | 89 (4.2) □ | 170 (3.6) □ |
| Level of knowledge, mean (SD) | 0.8 (1.4) | 0.4 (1.1) | 0.6 (1.3) |
| **Biological mother alive** |  |  |  |
| Yes, n (%) | 2,218 (87.3) □ | 1,881 (88.1) □ | 4,099 (87.7) □ |
| No, n (%) | 141 (5.6) □ | 104 (4.9) □ | 245 (5.2) □ |
| Do not know, n (%) | 5 (0.2) □ | 1 (0.0) □ | 6 (0.1) □ |
| Not declared, n (%) | 176 (6.9) □ | 149 (7.0) □ | 325 (7.0) □ |
| **Biological father alive** □ |  |  |  |
| Yes, n (%) □ | 2,086 (82.1) □ | 1,751 (82.0) □ | 3,837 (82.1) □ |
| No, n (%) □ | 257 (10.1) □ | 218 (10.2) | 475 (10.2) □ |
| Do not know, n (%) □ | 21 (0.8) □ | 17 (0.8) | 38 (0.8) □ |
| Not declared, n (%) | 176 (6.9) | 149 (7.0) | 325 (6.9) |
| **Residence** |  |  |  |
| Rural, n (%) | 1,617 (63.7) | 1,309 (61.3) | 2,926 (62.6) |
| Urban, n (%) | 923 (36.3) | 826 (38.7) | 1,749 (37.4) |
| **Strata** |  |  |  |
| Equateur, n (%) | 286 (11.3) | 241 (11.3) | 527 (11.3) |
| Kasai, n (%) | 545 (21.5) | 373 (17.5) | 918 (19.6) |
| Katanga, n (%) | 419 (16.5) | 316 (14.8) | 735 (15.7) |
| Kivu, n (%) | 417 (16.4) | 425 (19.9) | 842 (18.0) |
| Leopoldville, n (%) | 556 (21.9) | 533 (25.0) | 1,089 (23.3) |
| Oriental, n (%) | 317 (12.5) | 247 (11.6) | 564 (12.1) |

The overall prevalence of ever using any tobacco or nicotine product among adolescents was 18.53% (95% CI: 10.96 - 29.59) (Figure 2), equivalent to approximately 5.08 million adolescents out of the estimated adolescent population of 27.5 million. The prevalence of ever using tobacco products was 18.39% (95% CI: 10.90 - 29.35; ∼5.06 million adolescents in the general population), whilst that for nicotine products was 0.8% (95% CI: 0.19 - 3.36; ∼220,000 adolescents in the general population).

**Figure 2:**
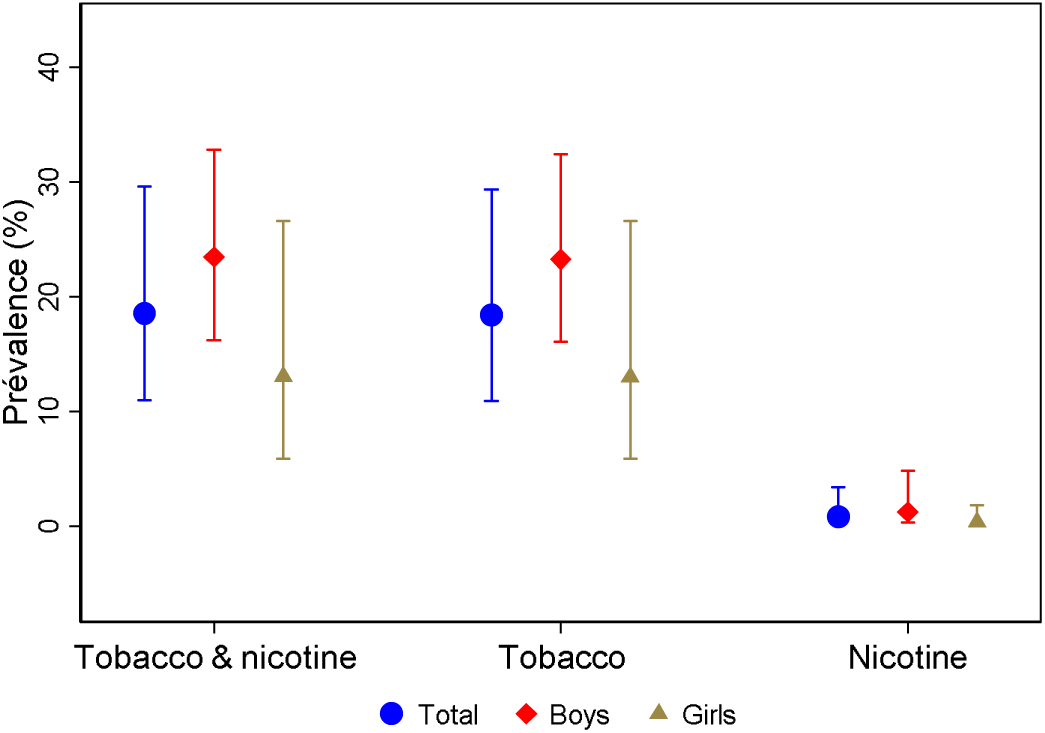
Prevalence of ever using any tobacco or nicotine products among adolescents.

Smoked tobacco products had an overall ever use prevalence of 12.28% (95% CI: 6.88 - 20.95; ∼3.38 million of general population adolescents) (Figure 3). Manufactured cigarettes were the most commonly ever used smoked tobacco products, with a prevalence of 9.1% (95% CI: 4.77 - 16.54; ∼2.50 million general population adolescents), followed by roll-your-own cigarettes at 5.3% (95% CI: 2.60 - 10.57; ∼1.46 million general population adolescents) and shisha at 2.2% (95% CI: 1.26 - 3.92; ∼605,000 general population adolescents). The overall prevalence of ever smoking cigarettes, either manufactured and roll-your-own, was 11.3% (95% CI: 6.34 - 19.34; ∼ 3.11 million general population adolescents).

**Figure 3:**
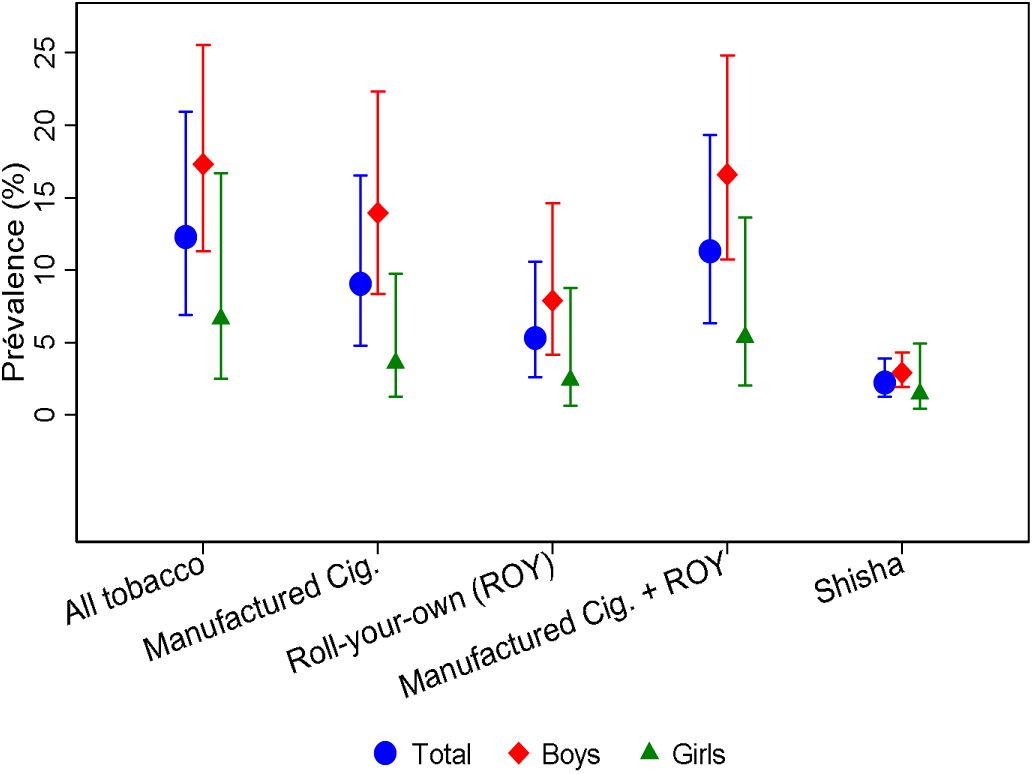
Prevalence of ever using smoked tobacco products among adolescents.

Smokeless tobacco products had an ever use prevalence of 9.00% (95% CI: 5.60 - 14.14; ∼2.48 million general population adolescents), whilst this was 0.2% (95% CI: 0.06 - 0.51; ∼55,000 general population adolescents) for heated tobacco products (Figure 4). Ever use prevalence of nicotine pouches was 0.66% (95% CI: 0.13 - 3.24; ∼192,000 general population adolescents) and that for electronic cigarettes was 0.15% (95% CI: 0.02 - 0.98; ∼55,000 population adolescents).

**Figure 4:**
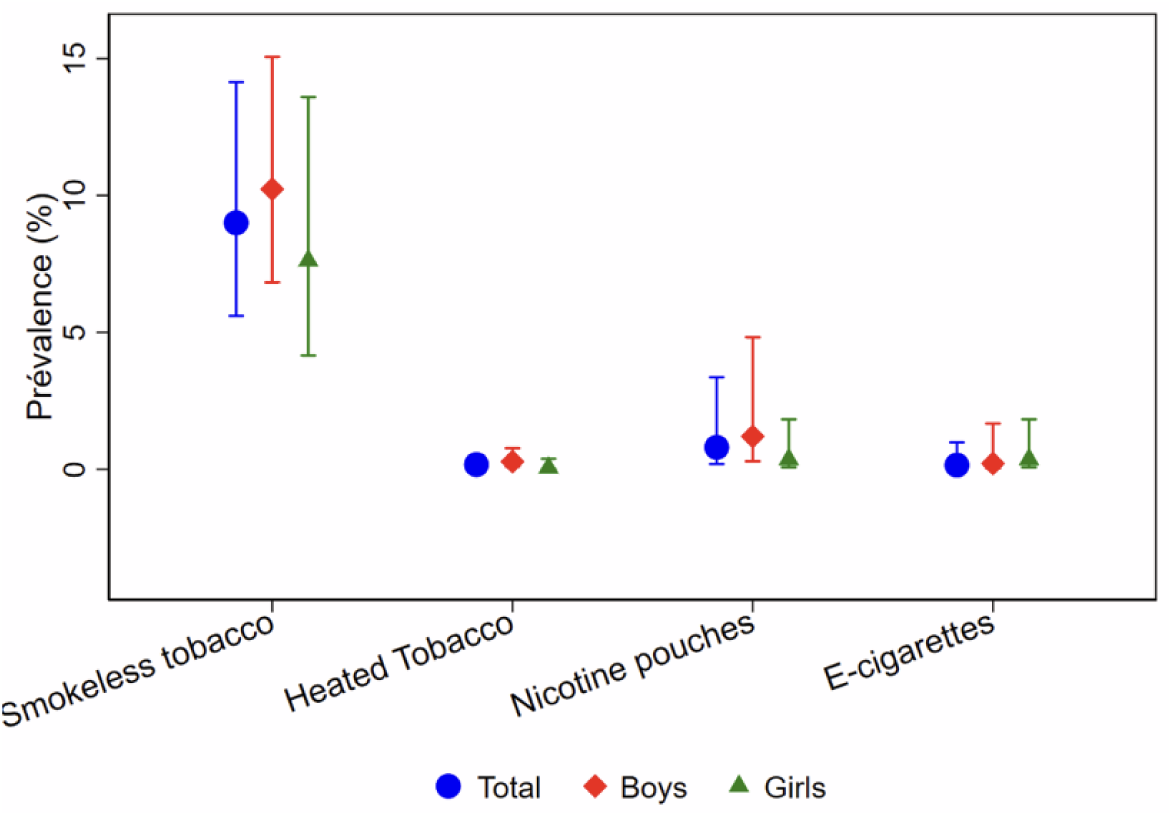
Prevalence of ever using smokeless tobacco, heated tobacco, nicotine pouches among adolescents.

As shown in Figures 2, 3 and 4, the prevalence of ever use was consistently higher among boys than girls, although the differences were not statistically significant. The results by age, schooling status, residence, wealth quintile, marital status, religion, functional disability and strata are also provided in Supplementary Material Tables 2 to 4.

### Current use prevalence

#### Any tobacco and nicotine products

The overall prevalence of the current use of any tobacco or nicotine product was 11.87% (95% CI: 6.93 - 19.58) which is approximately 3.27 million adolescents out of a total of 27.48 million in the general population (Table 2). This was almost entirely driven by tobacco product use with a current use prevalence of 11.85% (95% CI: 6.92 - 19.55, equating to approximately 3.24 million adolescents in the general population). The use of nicotine-only products was much lower than that of tobacco products, with a current use prevalence of 0.60% (95% CI: 0.10 - 3.40), which equates to ∼165 thousand adolescents in the general population. Table 2 also shows prevalence by sex, age, schooling status, residence, wealth quintile, marital status, religion, functional disability and strata. The current use prevalence was higher in boys than girls, higher age-groups, those out-of-school when compared to those who were in-school, and those residing in urban compared to those residing in rural areas. However, the differences were not statistically significant.

**Table 2:** The prevalence of current use of tobacco and nicotine products.

| **Characteristic** | **Tobacco and nicotine products**<br>**% (95% CI)** | **Tobacco products**<br>**% (95% CI)** | **Nicotine products**<br>**% (95% CI)** |
| --- | --- | --- | --- |
| **Overall** | **11.87 (6.93 - 19.58)** | **11.85 (6.92 - 19.55)** | **0.60 (0.10 - 3.40)** |
| **Sex** |  |  |  |
| Boys | 16.15 (9.93 - 25.17) | 16.13 (9.92 - 25.15) | 0.84 (0.15 - 4.64) |
| Girls | 7.09 (3.76 - 13.00) | 7.07 (3.74 - 12.97) | 0.33 (0.06 - 1.90) |
| **Age** |  |  |  |
| 10 - 12 yrs | 8.94 (4.96 - 15.59) | 8.91 (4.94 - 15.57) | 0.72 (0.09 - 5.71) |
| 13 -15 yrs | 11.8 (6.72 - 19.91) | 11.78 (6.70 - 19.87) | 0.28 (0.04 - 1.85) |
| 16 - 17yrs | 18.94 (10.18 - 32.51) | 18.94 (10.18 - 32.51) | 0.93 (0.23 - 3.70) |
| **Schooling status** |  |  |  |
| In-school | 11.24 (6.60 - 18.50) | 11.22 (6.58 - 18.47) | 0.51 (0.09 - 2.71) |
| Out-of-school | 16.38 (9.39 - 27.00) | 16.38 (9.39 - 27.00) | 1.26 (0.17 - 8.65) |
| **Residence** |  |  |  |
| Rural | 10.58 (5.93 - 18.17) | 10.58 (5.93 - 18.17) | 0.28 (0.03 - 2.39) |
| Urban | 16.98 (9.89 - 27.61) | 16.9 (9.84 - 27.47) | 1.84 (0.39 - 8.30) |
| **Wealth quintile** |  |  |  |
| 1st quintile: lowest | 11.33 (4.86 - 24.19) | 11.3 (4.85 - 24.13) | 0.73 (0.08 - 6.41) |
| 2nd quintile: low | 14.61 (10.07 - 20.72) | 14.58 (10.05 - 20.69) | 0.66 (0.08 - 4.98) |
| 3rd quintile: middle | 11.19 (5.26 - 22.22) | 11.15 (5.25 - 22.13) | 0.95 (0.15 - 5.74) |
| 4th quintile: high | 11 (5.01 - 22.47) | 11 (5.01 - 22.47) | 0.15 (0.02 - 0.96) |
| 5th quintile: highest | 11.19 (7.56 - 16.25) | 11.19 (7.56 - 16.25) | 0.46 (0.08 - 2.63) |
| **Marital status** |  |  |  |
| Not in a union | 17.18 (10.36 - 27.14) | 17.15 (10.34 - 27.10) | 0.65 (0.16 - 2.64) |
| In a union | 19.83 (6.33 - 47.49) | 19.83 (6.33 - 47.49) | 4.84 (0.99 - 20.65) |
| **Engagement in work** |  |  |  |
| Employee | 15.98 (9.56 - 25.48) | 15.98 (9.56 - 25.48) |  |
| Self-employed | 21.2 (7.00 - 49.04) | 21.2 (7.00 - 49.04) | 1.3 (0.25 - 6.40) |
| No work | 11.13 (6.25 - 19.02) | 11.1 (6.24 - 18.99) | 0.57 (0.07 - 4.25) |
| **Religion** |  |  |  |
| No religion | 29.88 (18.80 - 43.95) | 29.88 (18.80 - 43.95) | 5.13 (1.00 - 22.50) |
| Christianity | 10.99 (6.11 - 18.98) | 10.97 (6.09 - 18.95) | 0.33 (0.07 - 1.50) |
| Islam | 15.11 (4.53 - 40.03) | 15.11 (4.53 - 40.03) | 3.11 (0.37 - 21.55) |
| Hinduism | 37.16 (25.93 - 49.97) | 37.16 (25.93 - 49.97) |  |
| Other | 8.42 (2.88 - 22.19) | 8.42 (2.88 - 22.19) |  |
| **Functional disability** |  |  |  |
| Person with disability | 10.55 (6.08 - 17.68) | 10.55 (6.08 - 17.68) |  |
| Person without disability | 11.94 (6.66 - 20.48) | 11.92 (6.65 - 20.45) | 0.63 (0.11 - 3.57) |
| **Strata** |  |  |  |
| Equateur | 12.65 (6.86 - 22.17) | 12.36 (6.82 - 21.38) | 0.56 (0.18 - 1.74) |
| Kasai | 24.06 (11.04 - 44.73) | 24.06 (11.04 - 44.73) | 3.42 (0.54 - 18.85) |
| Katanga | 17.06 (3.60 - 53.11) | 17.06 (3.60 - 53.11) | 0.09 (0.01 - 0.90) |
| Kivu | 4.37 (1.01 - 16.95) | 4.37 (1.01 - 16.95) | 0.01 (0.00 - 0.04) |
| Leopoldville | 11.13 (6.44 - 18.55) | 11.13 (6.44 - 18.55) |  |
| Oriental | 30.05 (27.30 - 32.94) | 29.93 (27.01 - 33.03) | 1.18 (0.22 - 5.95) |

#### Smoked tobacco products

The overall prevalence of current any smoked tobacco product use among adolescents was 7.98% (95% CI: 4.23 - 14.55), corresponding to ∼ 2.19 million adolescents. This was primarily driven by current cigarette smoking, which had a prevalence of 7.41% (95% CI: 3.89 - 13.66) which represents about 2.04 million adolescents in the general population. Manufactured cigarettes had a current use prevalence of 6.65% (95% CI: 3.61 - 11.93), representing roughly 1.83 million adolescents in the general population, and that of roll-your-own cigarettes was 3.31% (95% CI: 1.31 - 8.13), equating to about 910 thousand general population adolescents. 1.37% (95% CI: 0.76 - 2.45) of adolescents reported that they were current shisha users, and this equates to approximately 380 thousand general population adolescents (Table 3).

**Table 3:**
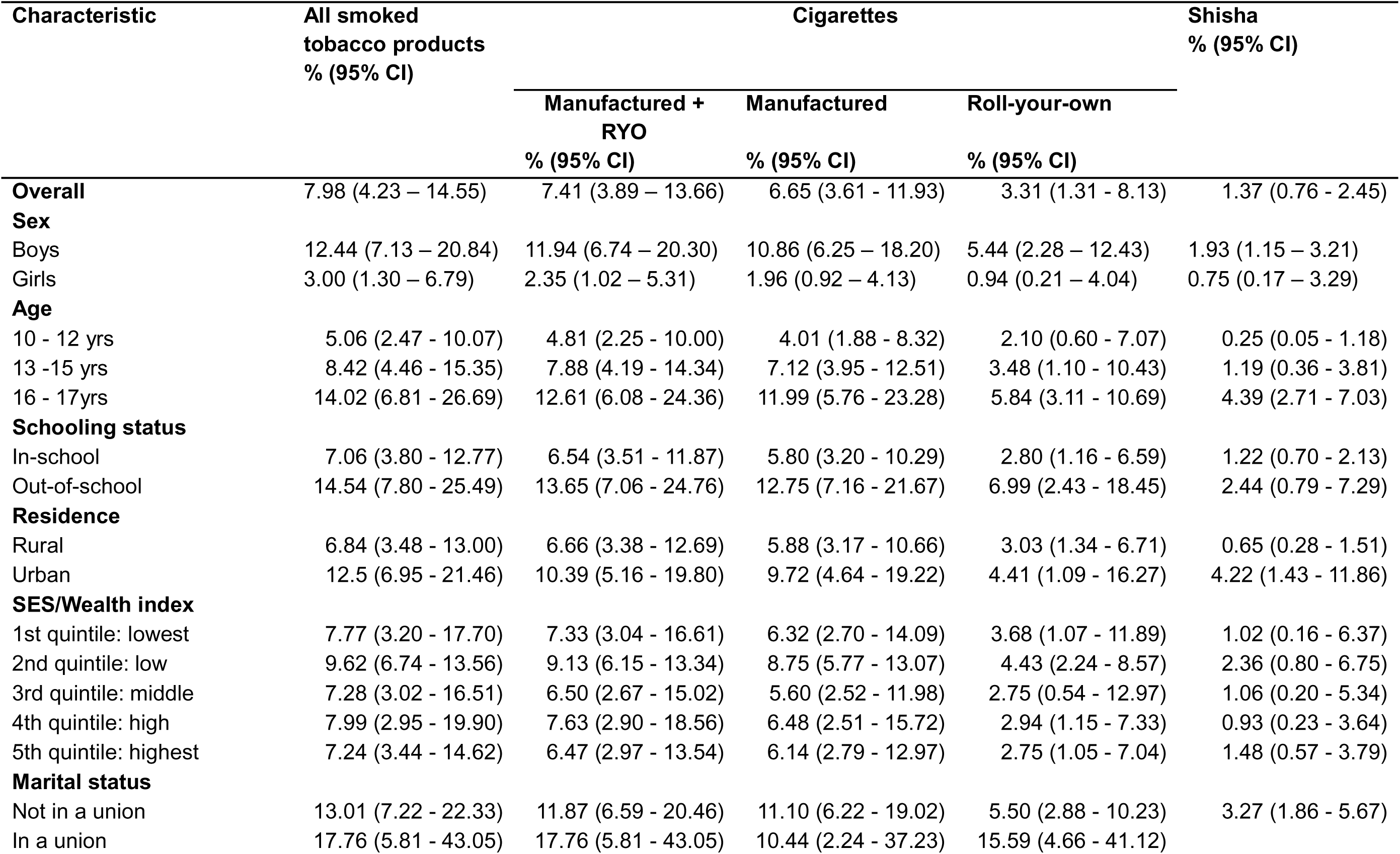
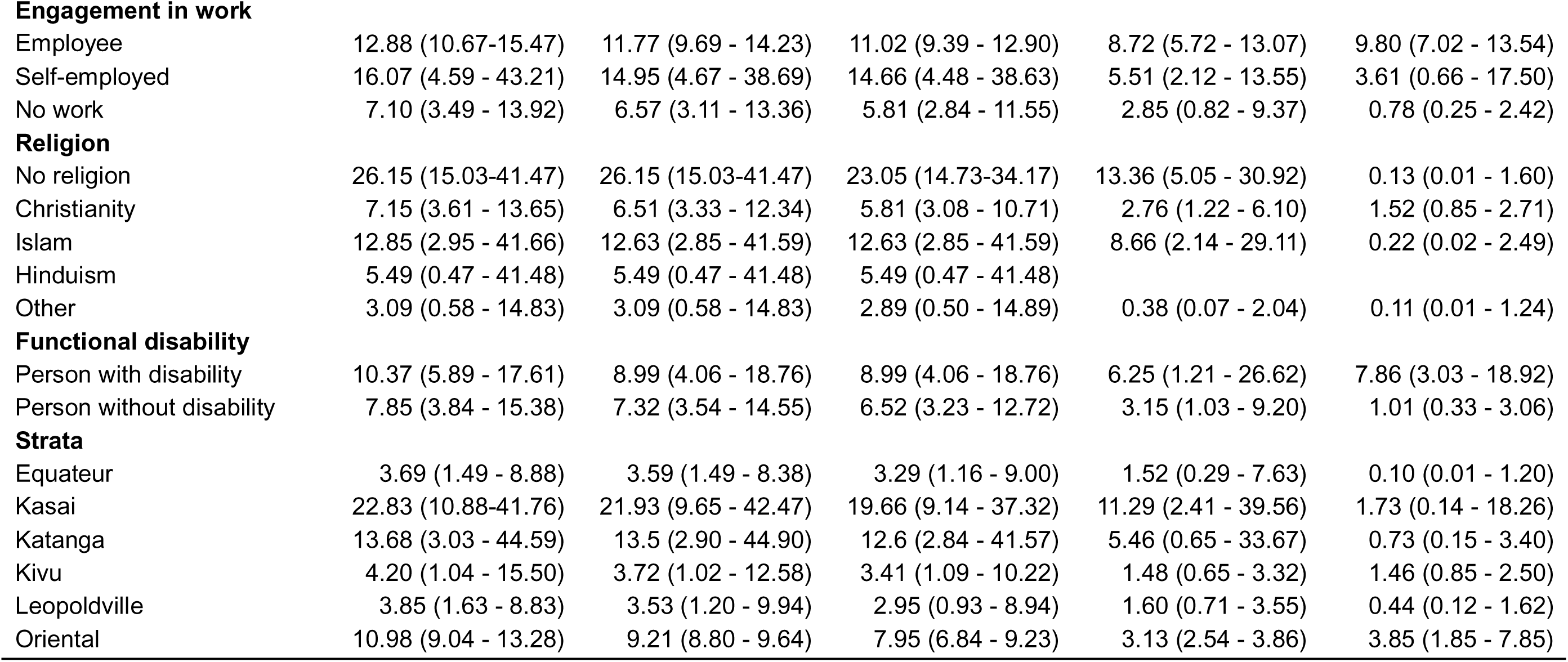
The prevalence of current use of smoked tobacco products.

| Characteristic | All smoked tobacco products<br>% (95% CI) | Cigarettes |  |  | Shisha<br>% (95% CI) |
| --- | --- | --- | --- | --- | --- |
|  |  | Manufactured + RYO<br>% (95% CI) | Manufactured<br>% (95% CI) | Roll-your-own<br>% (95% CI) |  |
| **Overall** | 7.98 (4.23 – 14.55) | 7.41 (3.89 – 13.66) | 6.65 (3.61 - 11.93) | 3.31 (1.31 - 8.13) | 1.37 (0.76 - 2.45) |
| **Sex** |  |  |  |  |  |
| Boys | 12.44 (7.13 – 20.84) | 11.94 (6.74 – 20.30) | 10.86 (6.25 – 18.20) | 5.44 (2.28 – 12.43) | 1.93 (1.15 – 3.21) |
| Girls | 3.00 (1.30 – 6.79) | 2.35 (1.02 – 5.31) | 1.96 (0.92 – 4.13) | 0.94 (0.21 – 4.04) | 0.75 (0.17 – 3.29) |
| **Age** |  |  |  |  |  |
| 10 - 12 yrs | 5.06 (2.47 - 10.07) | 4.81 (2.25 - 10.00) | 4.01 (1.88 - 8.32) | 2.10 (0.60 - 7.07) | 0.25 (0.05 - 1.18) |
| 13 -15 yrs | 8.42 (4.46 - 15.35) | 7.88 (4.19 - 14.34) | 7.12 (3.95 - 12.51) | 3.48 (1.10 - 10.43) | 1.19 (0.36 - 3.81) |
| 16 - 17yrs | 14.02 (6.81 - 26.69) | 12.61 (6.08 - 24.36) | 11.99 (5.76 - 23.28) | 5.84 (3.11 - 10.69) | 4.39 (2.71 - 7.03) |
| **Schooling status** |  |  |  |  |  |
| In-school | 7.06 (3.80 - 12.77) | 6.54 (3.51 - 11.87) | 5.80 (3.20 - 10.29) | 2.80 (1.16 - 6.59) | 1.22 (0.70 - 2.13) |
| Out-of-school | 14.54 (7.80 - 25.49) | 13.65 (7.06 - 24.76) | 12.75 (7.16 - 21.67) | 6.99 (2.43 - 18.45) | 2.44 (0.79 - 7.29) |
| **Residence** |  |  |  |  |  |
| Rural | 6.84 (3.48 - 13.00) | 6.66 (3.38 - 12.69) | 5.88 (3.17 - 10.66) | 3.03 (1.34 - 6.71) | 0.65 (0.28 - 1.51) |
| Urban | 12.5 (6.95 - 21.46) | 10.39 (5.16 - 19.80) | 9.72 (4.64 - 19.22) | 4.41 (1.09 - 16.27) | 4.22 (1.43 - 11.86) |
| **SES/Wealth index** |  |  |  |  |  |
| 1st quintile: lowest | 7.77 (3.20 - 17.70) | 7.33 (3.04 - 16.61) | 6.32 (2.70 - 14.09) | 3.68 (1.07 - 11.89) | 1.02 (0.16 - 6.37) |
| 2nd quintile: low | 9.62 (6.74 - 13.56) | 9.13 (6.15 - 13.34) | 8.75 (5.77 - 13.07) | 4.43 (2.24 - 8.57) | 2.36 (0.80 - 6.75) |
| 3rd quintile: middle | 7.28 (3.02 - 16.51) | 6.50 (2.67 - 15.02) | 5.60 (2.52 - 11.98) | 2.75 (0.54 - 12.97) | 1.06 (0.20 - 5.34) |
| 4th quintile: high | 7.99 (2.95 - 19.90) | 7.63 (2.90 - 18.56) | 6.48 (2.51 - 15.72) | 2.94 (1.15 - 7.33) | 0.93 (0.23 - 3.64) |
| 5th quintile: highest | 7.24 (3.44 - 14.62) | 6.47 (2.97 - 13.54) | 6.14 (2.79 - 12.97) | 2.75 (1.05 - 7.04) | 1.48 (0.57 - 3.79) |
| **Marital status** |  |  |  |  |  |
| Not in a union | 13.01 (7.22 - 22.33) | 11.87 (6.59 - 20.46) | 11.10 (6.22 - 19.02) | 5.50 (2.88 - 10.23) | 3.27 (1.86 - 5.67) |
| In a union | 17.76 (5.81 - 43.05) | 17.76 (5.81 - 43.05) | 10.44 (2.24 - 37.23) | 15.59 (4.66 - 41.12) |  |
| **Engagement in work** |  |  |  |  |  |
| Employee | 12.88 (10.67-15.47) | 11.77 (9.69 - 14.23) | 11.02 (9.39 - 12.90) | 8.72 (5.72 - 13.07) | 9.80 (7.02 - 13.54) |
| Self-employed | 16.07 (4.59 - 43.21) | 14.95 (4.67 - 38.69) | 14.66 (4.48 - 38.63) | 5.51 (2.12 - 13.55) | 3.61 (0.66 - 17.50) |
| No work | 7.10 (3.49 - 13.92) | 6.57 (3.11 - 13.36) | 5.81 (2.84 - 11.55) | 2.85 (0.82 - 9.37) | 0.78 (0.25 - 2.42) |
| **Religion** |  |  |  |  |  |
| No religion | 26.15 (15.03-41.47) | 26.15 (15.03-41.47) | 23.05 (14.73-34.17) | 13.36 (5.05 - 30.92) | 0.13 (0.01 - 1.60) |
| Christianity | 7.15 (3.61 - 13.65) | 6.51 (3.33 - 12.34) | 5.81 (3.08 - 10.71) | 2.76 (1.22 - 6.10) | 1.52 (0.85 - 2.71) |
| Islam | 12.85 (2.95 - 41.66) | 12.63 (2.85 - 41.59) | 12.63 (2.85 - 41.59) | 8.66 (2.14 - 29.11) | 0.22 (0.02 - 2.49) |
| Hinduism | 5.49 (0.47 - 41.48) | 5.49 (0.47 - 41.48) | 5.49 (0.47 - 41.48) |  |  |
| Other | 3.09 (0.58 - 14.83) | 3.09 (0.58 - 14.83) | 2.89 (0.50 - 14.89) | 0.38 (0.07 - 2.04) | 0.11 (0.01 - 1.24) |
| **Functional disability** |  |  |  |  |  |
| Person with disability | 10.37 (5.89 - 17.61) | 8.99 (4.06 - 18.76) | 8.99 (4.06 - 18.76) | 6.25 (1.21 - 26.62) | 7.86 (3.03 - 18.92) |
| Person without disability | 7.85 (3.84 - 15.38) | 7.32 (3.54 - 14.55) | 6.52 (3.23 - 12.72) | 3.15 (1.03 - 9.20) | 1.01 (0.33 - 3.06) |
| **Strata** |  |  |  |  |  |
| Equateur | 3.69 (1.49 - 8.88) | 3.59 (1.49 - 8.38) | 3.29 (1.16 - 9.00) | 1.52 (0.29 - 7.63) | 0.10 (0.01 - 1.20) |
| Kasai | 22.83 (10.88-41.76) | 21.93 (9.65 - 42.47) | 19.66 (9.14 - 37.32) | 11.29 (2.41 - 39.56) | 1.73 (0.14 - 18.26) |
| Katanga | 13.68 (3.03 - 44.59) | 13.5 (2.90 - 44.90) | 12.6 (2.84 - 41.57) | 5.46 (0.65 - 33.67) | 0.73 (0.15 - 3.40) |
| Kivu | 4.20 (1.04 - 15.50) | 3.72 (1.02 - 12.58) | 3.41 (1.09 - 10.22) | 1.48 (0.65 - 3.32) | 1.46 (0.85 - 2.50) |
| Leopoldville | 3.85 (1.63 - 8.83) | 3.53 (1.20 - 9.94) | 2.95 (0.93 - 8.94) | 1.60 (0.71 - 3.55) | 0.44 (0.12 - 1.62) |
| Oriental | 10.98 (9.04 - 13.28) | 9.21 (8.80 - 9.64) | 7.95 (6.84 - 9.23) | 3.13 (2.54 - 3.86) | 3.85 (1.85 - 7.85) |

**Table 4:**
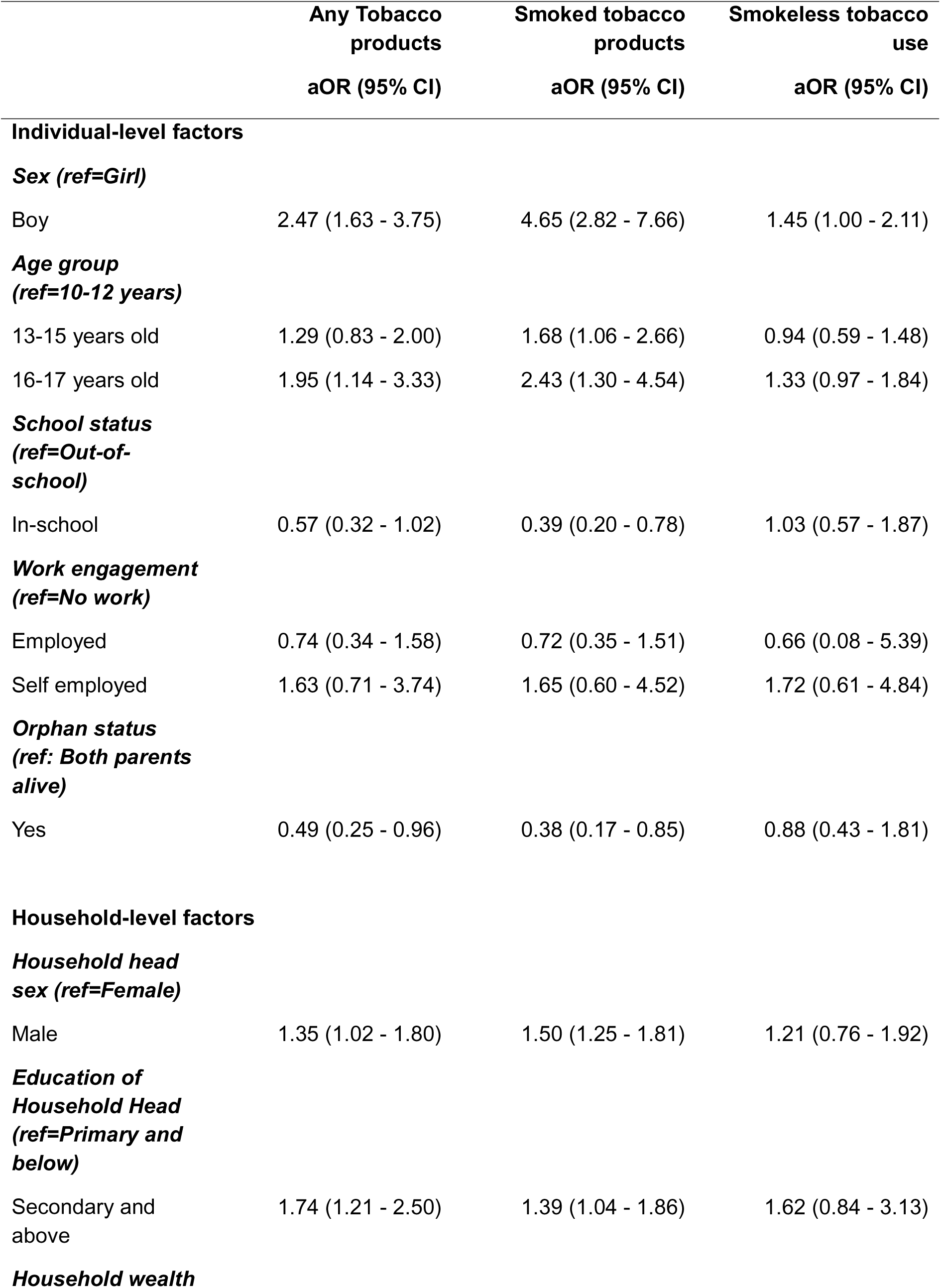
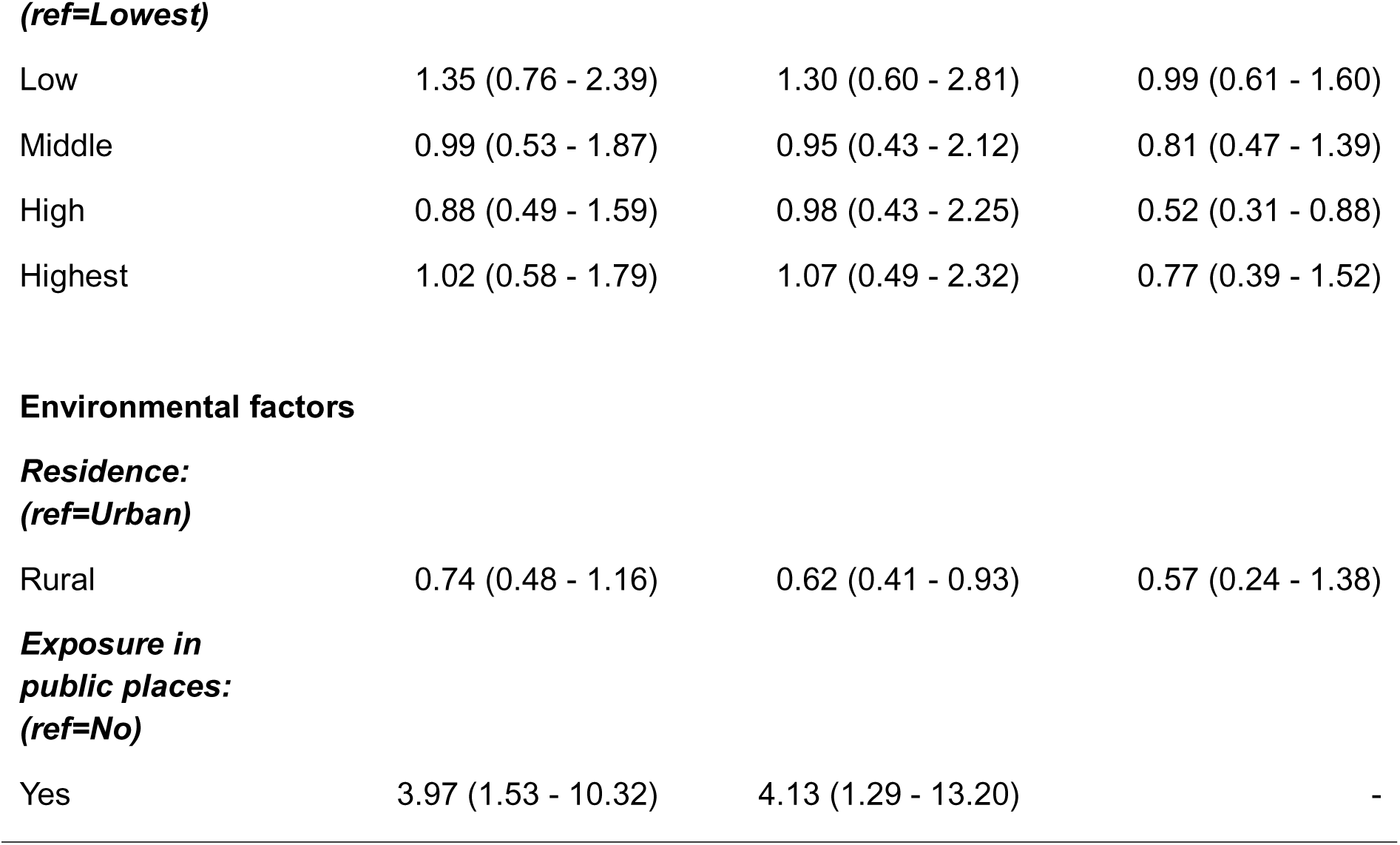
Factors associated with current use of tobacco and nicotine products.

Across all product categories, the current use prevalence was consistently higher in boys than girls, higher age-groups, those out-of-school when compared to those who were in-school, and those residing in urban compared to those residing in rural areas (Table 3). However, the differences were only statistically significant for all smoked products and cigarette smoking between boys and girls. Table 3 also shows prevalence by wealth quintile, marital status, religion, functional disability and strata.

#### Heated tobacco products (HTP)

The current use of HTP was 0.11% (95% CI: 0.11 - 0.11), and it was reported exclusively among boys. This equates to about 30,228 adolescent boys in the general population. The results by sex, age, schooling status, residence, wealth quintile, marital status, religion, functional disability and strata are provided in Supplementary Material Table 5.

#### Smokeless tobacco products

The overall current use prevalence was 5.86% (95% CI: 3.42 - 9.87) for smokeless tobacco products, representing ∼ 1.61 million general population adolescents (Supplementary Material Table 6). Current use prevalence was higher among boys (6.71%) than girls (4.91%), those living in urban areas as compared to rural areas, and among adolescents that did not report any functional disability than those that reported at least one. However, these differences were not statistically significant. The current use prevalence by age, schooling status, wealth quintile, marital status, religion and strata are also provided in Supplementary Material Table 6.

#### Nicotine products

The overall prevalence of current use of nicotine products (i.e., electronic cigarettes and nicotine pouches) was 0.60% (95% CI: 0.10 - 3.40) (Supplementary Material Table 7). This translates to about ∼ 165 thousand adolescents in the general population. This was mainly due to the use of nicotine pouches where 0.56% (CI: 95% 0.09 - 3.55) of the adolescents reported being current users, which translates to about 154 thousand adolescents in the general population. Current use of electronic cigarettes was reported by 0.04 % (CI 0.01 - 0.16) of the adolescents, which equates to about 11 thousand adolescents in the general population. Results by sex, age, schooling status, residence, wealth quintile, marital status, religion, functional disability and strata are also provided in Supplementary Material Table 7.

### Factors associated with tobacco product use

#### Any tobacco use

Boys were significantly more likely to use tobacco products compared to girls (aOR = 2.47; 95% CI: 1.63 - 3.75) (Table 4). Older adolescents aged 16–17 years had higher odds of tobacco use compared to those aged 10–12 years (aOR = 1.95; 95% CI: 1.14 - 3.33). In-school adolescents were less likely to use tobacco than those out of school (aOR = 0.57; 95% CI: 0.32 - 1.02). Adolescents who had lost one or both parents were less likely to use tobacco products compared to those with both parents alive (aOR = 0.49; 95% CI: 0.25 - 0.96). Living in male-headed households was associated with higher odds of tobacco use compared to female-headed households (aOR = 1.35; 95% CI: 1.02 - 1.80). Adolescents from households where the head had at least secondary education were more likely to use tobacco (aOR = 1.74; 95% CI: 1.21 - 2.50). Exposure to tobacco use in public places was strongly associated with tobacco use (aOR = 3.97; 95% CI: 1.53 - 10.32).

#### Smoked tobacco products

Boys had significantly higher odds of smoking tobacco compared to girls (aOR = 4.65; 95% CI: 2.82 - 7.66) (Table 4). Adolescents aged 13–15 years (aOR = 1.68; 95% CI: 1.06 - 2.66) and 16–17 years (aOR = 2.43; 95% CI: 1.30 - 4.54) were more likely to smoke than those aged 10–12 years. In-school adolescents were less likely to smoke compared to those out of school (aOR = 0.39; 95% CI: 0.20 - 0.78). Adolescents who had lost one or both parents were also less likely to smoke (aOR = 0.38; 95% CI: 0.17 - 0.85). Living in male-headed households was associated with higher odds of smoking (aOR = 1.50; 95% CI: 1.25 - 1.81), as did having a household head with at least secondary education (aOR = 1.39; 95% CI: 1.04 - 1.86). Adolescents living in rural areas were less likely to smoke compared to those in urban areas (aOR = 0.62; 95% CI: 0.41 - 0.93). Exposure to tobacco use in public places was also associated with higher odds of smoking (aOR = 4.13; 95% CI: 1.29 - 13.20).

#### Smokeless tobacco products

Boys were more likely to use smokeless tobacco than girls (aOR = 1.45; 95% CI: 1.00 - 2.11) (Table 4). Adolescents aged 16–17 years showed a marginally higher likelihood of use compared to those aged 10–12 years (aOR = 1.33; 95% CI: 0.97 - 1.84). School attendance and orphan status were not significantly associated with smokeless tobacco use. Adolescents from wealthier households (high wealth category) were less likely to use smokeless tobacco compared to those from the lowest wealth group (aOR = 0.52; 95% CI: 0.31 - 0.88). No significant associations were observed for residence or most other household-level factors.

## Discussion

To our knowledge, this study is the first of its kind in the DRC. The prevalence of ever and current use were high for tobacco or nicotine products (18.53% and 11.87%, respectively), tobacco products (18.39% and 11.85%, respectively), smoked tobacco products (12.28% and 7.98%, respectively), and smokeless tobacco (9.00% and 5.86%, respectively). Current use of smoked tobacco was mainly manufactured and RYO cigarettes (7.41%), with a small proportion reporting shisha use (1.37%). In addition, only small proportions of adolescents had ever used or were currently using nicotine products (i.e., pouches and electronic cigarettes; 0.56% and 0.04%, respectively).

Our results for ever or current use of cigarettes were in line with those reported in the DRC GYTS study in Lubumbashi and Kinshasa among 13–15-year-olds in school (19.0 and 19.5% for ever smoking, and 7.5% and 8.2% for current smoking, respectively) [11]. However, proportions who reported ever or currently using any tobacco products were much higher in the DRC GYTS study: 28.0% and 36.6% for ever smoking, and 24.8% and 28.8% for current smoking, for Lubumbashi and Kinshasa respectively. The difference could be due to differences in the definition of tobacco products and how this was measured. The overall prevalence of current tobacco use, cigarette smoking and smokeless tobacco use found in our study were also consistent with recently reported pooled prevalence estimates for African countries from GYTS and GSHS data [21, 22]. However, studies in Kenya and Nigeria that were conducted in the same year using the same methodology as our study reported much lower prevalence estimates: e.g., for tobacco products this was 2.5% in Kenya [23] and 3.6% in Nigeria [24]. This might be reflective of the more comprehensive tobacco control policies and higher levels of implementation in Kenya and Nigeria, when compared to the DRC. Although we observed low prevalence of use of shisha, heated tobacco products, electronic cigarettes and nicotine pouches, findings from other studies indicate a rise in the use of these products in the African region, with some countries reacting by implementing complete bans [25].

The well-established gender difference in tobacco use, particularly smoked tobacco [26] was also observed in our study, with girls being less likely to be current users of smoked tobacco products than boys. In addition, school attendance appeared to be a protective factor against tobacco use, which might reflect higher access to health-related information and supervision, and reduced exposure to peer-driven experimentation for those who are a structured school environment when compared to those who are out-of-school [27]. Our observation that those who lived in female-headed households were less likely to use tobacco compared to those from male-headed households might reflect a higher prevalence of smoking among men generally. This might mean those in male headed households are more likely to be exposed to smoking in their households. It is well established that adolescents who have family members who use tobacco are more likely to be current tobacco users than those who do not have family members who use tobacco, highlighting the role of social modelling. Access to pocket money was associated with increased tobacco use, as reported elsewhere [28], whereas the protective effect observed among adolescents with functional disabilities may reflect reduced exposure or access to the products, although this requires further investigation.

### Strengths and limitations

We had high study enrollment rates with 99% and 96% of sampled households and adolescents, respectively, completing the study surveys. It took 12 weeks to enroll the required sample size, which is in line with the recommended timescales for other household-based surveys [14]. Data completeness was also high (96%). These figures demonstrate that collecting data on tobacco and nicotine products among adolescents using household survey is feasible and acceptable. We included a broader age range from 10 to 17 years, and out-of-school adolescents who are normally excluded in most tobacco use studies. The proportion of out-of-school adolescents in our sample was 11.5%, which is ∼3.16 million adolescents in the general population. Our study also provides new data on the use of heated tobacco products and nicotine products: this data is lacking in the Sub-Saharan African context. We also provided data on the exploring individual-, household- and environmental-level factors associated with tobacco use among adolescents. Nevertheless, some adolescents, particularly girls, might have been reluctant to self-report tobacco and nicotine product use, behaviours that might be viewed as socially undesirable. This might have led to under-reporting despite our efforts to ensure privacy and confidentiality.

### Policy implications

Our study demonstrates that the prevalence of tobacco use among adolescents in the DRC is high. There is a need to strengthen policies that make tobacco and nicotine products less accessible and less appealing to this group. This could include making the products less affordable by raising excise taxes so that they are at least 75% of the retail price instead of the current 38.7%. Young people are very sensitive to price increases, which makes raising taxes an effective way of reducing consumption in this population [29]. The DRC should also ban the sale of single cigarette sticks or small packets because it undermines tax policies by enabling those who cannot afford cigarette packs to switch to sticks rather than quit. In addition, when young people buy sticks, they do not see health warnings, and, therefore, do not benefit from the deterrent effects of these warnings [30]. Tobacco control efforts in the DRC could also be strengthened by banning the sale of tobacco and nicotine products in or near locations that are frequented by children and adolescents such as educational facilities and playgrounds, point of sale product displays, and the sale of flavoured tobacco and nicotine products. Tobacco control should not only address current products but also anticipate industry product innovation and loopholes that may weaken existing regulations. There is also a need to enhance tobacco cessation support for adolescents.

### Research implications

Future research should consider regular surveillance of tobacco and nicotine product use using both quantitative and qualitative study designs that enable prevalence estimates, the evaluation of the effectiveness of tobacco control policies and interventions and offer an in-depth understanding of the context within which use occurs. Longitudinal studies could also offer an understanding of how tobacco and nicotine product use behaviour develops, including the factors that influence transition from experimentation to continued use, and evaluate the long-term consequences of the behaviour.

## Supporting information

STROBE checklist

## Data Availability

All data produced in the present study are available upon reasonable request to the authors

## Ethical Considerations

The study was approved by the National Health Ethics Committee of the DRC (approval no. 513/CNES/BN/PMMF/2024, issued on 17 February 2024). Research permits and letters of support were obtained from the Ministry of Health and relevant provincial and local authorities. Participation was voluntary, and confidentiality and participant rights were safeguarded throughout. Due to cultural sensitivities around signing documents, verbal informed consent was obtained from adults and legal guardians, and verbal assent from adolescents, in accordance with approved protocols. Assent from adolescents was obtained in the presence of a household representative, and all consents were appropriately documented. Study information was provided in participants’ preferred language (French, Lingala, Swahili, Tshiluba, or Kikongo), with opportunities for clarification.

Data access was password-protected and restricted to key study personnel, ensuring data integrity and security. Each participant was assigned a unique study identifier, and all personal identifiers were removed prior to data analysis to safeguard confidentiality.

## Author contributions

NDM, CT, DMM, JDT, RP, TGA, and EK conceptualised the study, and funding was acquired by NDM, CT, DMM and EK. All authors contributed to the study design and methodology. The following curated and analysed the data: BWNN, CT and DMM. All authors contributed to the validation of results. Study supervision was by CT, BWNN and DMM, with oversight from NDM. The original draft of the manuscript was written by CT, BWNN, DMM and NDM, and all authors reviewed, edited and approved the final manuscript.

## Acknowledgments

We extend our deepest gratitude to the Ministère de la Santé Publique, Hygiène et Prévoyance Sociale of the Democratic Republic of Congo and, specifically, Programme National de Lutte Contre la Toxicomanie et les Substances Toxiques for their exceptional commitment to the realisation of this study. We are also sincerely thankful to the families and adolescents who generously shared their time and perspectives, as well as to the field investigators whose professionalism and dedication were essential to the successful completion of the research. Our gratitude also goes to the members of the study’s Advisory Board and Steering Committee for their invaluable guidance and support throughout the research process, as well as the Development Gateway team for their technical and coordination support.

## Reference

[1] World Health Organization (WHO). Tobacco fact sheet. 2025. https://www.who.int/docs/default-source/campaigns-and-initiatives/world-no-tobacco-day-2020/wntd-tobacco-fact-sheet.pdf.

[2] World Health Organization (WHO). WHO global report on trends in prevalence of tobacco use 2000–2030. 2024. https://brasil.un.org/sites/default/files/2024-01/OMS_Relatorio-Tendencias-Tabaco_2010_2030-eng.pdf.

[3] United Nations Development Programme. Annuaire statistique RDC. 2020. https://files.acquia.undp.org/public/migration/cd/UNDP-CD-ANNUAIRE-STAT.-2020-.pdf.

[4] Brown JL, Rosen D, Carmona MG, Parra N, Hurley M, Cohen JE. Spinning a global web: tactics used by Big Tobacco to attract children at tobacco points-of-sale. Tobacco Control 2023; 32: 645–651. 10.1136/tobaccocontrol-2021-057095.

[5] Struik LL, Dow-Fleisner S, Belliveau M, Thompson D, Janke R. Tactics for drawing youth to vaping: content analysis of electronic cigarette advertisements. Journal of Medical Internet Research 2020; 22: e18943. 10.2196/18943.

[6] Munyanga Mukungo S. Plan stratégique national de lutte contre les maladies non transmissibles En République Démocratique du Congo. 2016. https://www.iccp-portal.org/sites/default/files/plans/COD_B3_PLAN%20STRATEGIQUE%20MULTISECTORIEL%20MNT%202016-2020.pdf.

[7] World Health Organization (WHO). Tobacco and nicotine industry tactics addict youth for life. https://www.who.int/news/item/23-05-2024-tobacco-and-nicotine-industry-tactics-addict-youth-for-life.

[8] James PB, Bah AJ, Kabba JA, Kassim SA, Dalinjong PA. Prevalence and correlates of current tobacco use and non-user susceptibility to using tobacco products among school-going adolescents in 22 African countries: a secondary analysis of the 2013-2018 global youth tobacco surveys. Archives Public Health 2022;80:121. 10.1186/s13690-022-00881-8.

[9] African Tobacco Control Alliance(ACTA). Country Profile Congo Democratic Republic. 2024. https://atca-africa.org/country-profiles-congo-democratic-republic.

[10] Campaign for Tobacco Free Kids. Tobacco Control Laws: Democratic Republic of the Congo. 2026. https://www.tobaccocontrollaws.org/legislation/democratic-republic-of-the-congo/advertising-promotion-sponsorship.

[11] Mbuyu Muteba R, Banza LNC. Le tabagisme en milieu scolaire en République Démocratique du Congo. 2008. https://portal-uat.who.int/fctcapps/sites/default/files/2023-04/Annex2_GYTS_report_2008.pdf.

[12] A E O Ogwell, A N Aström, O Haugejorden. Socio-demographic factors of pupils who use tobacco in randomly-selected primary schools in Nairobi province, Kenya. East African Medical Journal 2003;80:235–41. 10.4314/eamj.v80i5.8693.

[13] Olawole-Isaac A, Ogundipe O, Amoo EO, Adeloye D. Substance use among adolescents in sub-Saharan Africa: a systematic review and meta-analysis. South African Journal of Child Health 2018;12:79. 10.7196/sajch.2018.v12i2.1524.

[14] Kisia L, Mohamed SF, Kyule G, Tchoupe C, Abolarin O, Pokothoane R, et al. Use of tobacco and nicotine products among adolescents in Sub-Saharan Africa: protocol for a population-based multi-country household survey. Frontiers in Public Health 2025;13:1562352. 10.3389/fpubh.2025.1562352.

[15] United Nations (ed). Designing household survey samples: practical guidelines. New York, NY: United Nations. 2008.

[16] Mavuta Zalula C, Whari L Imani, L Ngimbi S, Ngoie NL, Tshiswaka SM, Luboya EK, et al. Pratiques alimentaires des nourrissons: Connaissances, attitudes et pratiques des mères d’une commune urbaine de la ville de Lubumbashi, République Démocratique du Congo. Revue de l’Infirmier Congolais 2018;2:109–116.

[17] Ntambue AM, Tshiala RN, Malonga FK, Ilunga TM, Kamonayi JM, Kazadi ST, et al. Utilisation des méthodes contraceptives modernes en République Démocratique du Congo: prévalence et barrières dans la zone de santé de Dibindi à Mbuji-Mayi. Pan African Medical Journal 2017;26:199. 10.11604/pamj.2017.26.199.10897.

[18] Dobility, Inc. SurveyCTO (Version 2.70). https://www.surveycto.com. 2024.

[19] Bronfenbrenner U. Toward an experimental ecology of human development. American Psychologist 1977;32:513–531. 10.1037/0003-066X.32.7.513.

[20] McLeroy KR, Bibeau D, Steckler A, Glanz, K. An ecological perspective on health promotion programs. 1988;15:351–377. 10.1177/109019818801500401.

[21] Pokothoane R, Argefa TG, Tsague JD, Mdege ND. What are the factors associated with alcohol, cigarette and marijuana use among adolescents in Africa? Evidence from the Global School-based Health Survey. BMJ Open 2025;15:e089096. 10.1136/bmjopen-2024-089096.

[22] Pokothoane R, Agerfa TG, Miderho CC, Mdege ND. Prevalence and determinants of tobacco use among school-going adolescents in 53 African countries: Evidence from the Global Youth Tobacco Survey. Addictive Behaviors Reports 2025;21:100581. 10.1016/j.abrep.2024.100581.

[23] Ministry of Health (MoH), Kenya National Bureau of Statistics (KNBS), African Population and Health Research Center (APHRC) and Development Gateway (DG). Tobacco and Nicotine Use Among Adolescents in Kenya: Findings from the DaYTA Survey, 2024. Nairobi: APHRC. 2024.

[24] Federal Ministry of Health (FMoH), National Population Commission (NPC), African Population and Health Research Center (APHRC), APIN Public Health Initiatives (APIN), Research and Communications Services Ltd (RCS), Development Gateway (DG). Tobacco and Nicotine Use Among Adolescents in Nigeria: Findings from the DaYTA Survey, 2024. 2025.

[25] World Health Organization. WHO report on the global tobacco epidemic, 2025: warning about the dangers of tobacco. Geneva: World Health Organization. 2025.

[26] INS. Enquête par grappes à indicateurs multiples, 2017-2018, rapport de résultats de l’enquête. Kinshasa, République Démocratique du Congo. 2019. file:///C:/Users/chris/Downloads/congo-democratic-republic-of-the-2017-18-mics-sfr_french.pdf.

[27] Chido-Amajuoyi OG, Osaghae I, Agaku IT, Chen B, Mantey DS. Exposure to school-based tobacco prevention interventions in low-income and middle-income countries and its association with psychosocial predictors of smoking among adolescents: a pooled cross-sectional analysis of Global Youth Tobacco Survey data from 38 countries. BMJ Open 2024; 14: e070749. 10.1136/bmjopen-2022-070749.

[28] Jawad M, Li W, Filippidis FT. Sociodemographic inequalities in cigarette, smokeless tobacco, waterpipe tobacco, and electronic cigarette use among adolescents aged 12-16 years in 114 countries: a cross-sectional analysis. Tobacco Induced Diesses 2024; 22:151. 10.18332/tid/191824.

[29] Filby S, van Walbeek C. Cigarette prices and smoking among youth in 16 African countries: evidence from the Global Youth Tobacco Survey. Nicotine and Tobacco Research, 2022;24:1218–27. 10.1093/ntr/ntac017.

[30] Drovandi A, Teague P-A, Glass B, Malau-Aduli B. Smoker perceptions of health warnings on cigarette packaging and cigarette sticks: a four-country study. Tobacco Induced Diseases 2019;17:23. 10.18332/tid/104753.

