## Supplementary Material Table 1 for "Prevalence and factors associated with tobacco and nicotine product use among adolescents in the Democratic Republic of the Congo: evidence from a cross-sectional national household survey"

**Supplementary Material Table 1: Household background characteristics**

| **Household characteristics, mean (SD)** |  |
| --- | --- |
| Number of people in household | 4.7 (2.1) |
| Number of eligible adolescents in the household | 1.9 (1.0) |
| Age of household head | 41.6 (14.9) |
| **Sex (household head), n (%)** | |
| Male | 3,203 (68.5) |
| Female | 1,472 (31.5) |
| **Disability status (household head), n (%)** | |
| Person with disability | 193 (4.3) |
| Person without disability | 4,330 (95.7) |
| **Main income activity (household head), n (%)** | |
| None | 752 (16.6) |
| Unestablished own business | 653 (14.4) |
| Established own business | 78 (1.7) |
| Informal casual | 601 (13.3) |
| Informal salaried | 249 (5.5) |
| Formal salaried | 515 (11.4) |
| Formal casual | 138 (3.1) |
| Agriculture | 1,538 (34.0) |
| **Marital status (household head), n (%)** | |
| Not in a union | 481 (10.3) |
| Married | 3,048 (65.4) |
| In a union | 363 (7.8) |
| Divorced | 85 (1.8) |
| Separated | 243 (5.2) |
| Widowed | 427 (9.2) |
| Refused to answer | 10 (0.2) |
| **Education level (household head), n (%)** | |
| No education | 511 (11.4) |
| Primary | 1,062 (23.6) |
| Secondary | 2,401 (53.4) |
| Technical/Vocational | 113 (2.5) |
| Higher | 413 (9.2) |
| **Household wealth index, n (%)** | |
| Lowest | 936 (20) |
| Low | 937 (20) |
| Middle | 933 (20) |
| High | 936 (20) |
| Highest | 933 (20) |
| **Residence, n (%)** | |
| Rural | 2,926 (62.6) |
| Urban | 1,749 (37.4) |
| **Head of household health insurance, n (%)** | |
| Yes | 319 (7.1) |
| No | 4,205 (92.9) |
| **Number of household members with insurance, mean (SD)** | 0.2 (0.9) |
| **Strata, n (%)** | |
| Equateur | 527 (11.3) |
| Kasai | 918 (19.6) |
| Katanga | 735 (15.7) |
| Kivu | 842 (18.0) |
| Leopoldville | 1,089 (23.3) |
| Oriental | 564 (12.1) |
