## Supplementary Material Table 2 for "Prevalence and factors associated with tobacco and nicotine product use among adolescents in the Democratic Republic of the Congo: evidence from a cross-sectional national household survey"

**Supplementary Material Table 2: Prevalence of ever use of tobacco and nicotine products**

| **Characteristic** | **Tobacco and nicotine products**  **% (95% CI)** | **Tobacco products**  **% (95% CI)** | **Nicotine products**  **% (95% CI)** |
| --- | --- | --- | --- |
| **Overall** | 18.53 [10.96 - 29.59] | 18.39 [10.90 - 29.35] | 0.80 [0.19 - 3.36] |
| **Sex** |  |  |  |
| Boys | 23.46 [16.16 – 32.77] | 23.22 [16.03- 32.40] | 1.20 [0.29 – 4.82] |
| Girls | 13.03 [5.84 – 26.59] | 13.01 [5.83 – 26.56] | 0.36 [0.07 – 1.82] |
| **Age** |  |  |  |
| 10 - 12 yrs | 14.3 [9.72 - 20.55] | 14.24 [9.68 - 20.45] | 0.8 [0.12 - 5.29] |
| 13 -15 yrs | 18.95 [10.01 - 32.96] | 18.69 [9.92 - 32.44] | 0.59 [0.14 - 2.42] |
| 16 - 17yrs | 27.72 [14.36 - 46.72 | 27.64 [14.30 - 46.65] | 1.24 [0.32 - 4.67] |
| **Schooling status** |  |  |  |
| **In-school** | 18.29 [10.50 - 29.92 | 18.13 [22.17 - 24.76] | 0.73 [0.51 - 1.04] |
| **Out-of-school** | 20,27 [13.60 - 29.11 | 20.27 [28.40 - 36.29] | 1.37 [0.87 - 3.19] |
| **Residence** |  |  |  |
| Rural | 15.46 [9.29 - 24.62] | 15.46 [9.29 - 24.61] | 0.29 [0.04 - 2.26] |
| Urban | 30.72 [17.98 - 47.29] | 30.05 [17.89 - 45.86] | 2.84 [0.78 - 9.80] |
| **Wealth quintile** |  |  |  |
| 1st quintile: lowest | 19.86 [11.97 - 31.12] | 19.77 [11.94 - 30.93] | 0.81 [0.11 - 5.78] |
| 2nd quintile: low | 20.91 [12.53 - 32.81] | 20.88 [12.50 - 32.76] | 0.74 [0.12 - 4.53] |
| 3rd quintile: middle | 17.89 [7.20 - 37.95] | 17.77 [7.17 - 37.65] | 1.06 [0.20 - 5.32] |
| 4th quintile: high | 17.17 [8.23 - 32.37] | 16.73 [8.00 - 31.71] | 0.9 [0.14 - 5.47] |
| 5th quintile: highest | 16.79 [12.30 - 22.50] | 16.79 [12.30 - 22.50] | 0.48 [0.09 - 2.59] |
| **Marital status** |  |  |  |
| Not in a union | 25.16 [14.03 - 40.92] | 24.83 [13.86 - 40.42] | 1.08 [0.25 - 4.48] |
| In a union | 32.45 [8.82 - 70.47] | 32.45 [8.82 - 70.47] | 4.84 [0.99 - 20.65] |
| **Engagement in work** |  |  |  |
| Employee | 31.59 [23.19 - 41.40] | 31.59 [23.19 - 41.40] | 1.02 [0.09 - 10.48] |
| Self-employed | 26.29 [8.75 - 57.01] | 26.26 [8.74 - 56.98] | 1.33 [0.27 - 6.32] |
| No work | 17.75 [9.54 - 30.62] | 17.58 [9.45 - 30.35] | 0.77 [0.16 - 3.61] |
| **Religion** |  |  |  |
| No religion | 36.6 [26.57 - 47.94] | 36.6 [26.57 - 47.94] | 5.13 [1.00 - 22.50] |
| Christianity | 17.77 [9.78 - 30.11] | 17.63 [9.72 - 29.84] | 0.54 [0.13 - 2.26] |
| Islam | 17.26 [6.46 - 38.64] | 16.88 [6.06 - 39.03] | 3.49 [0.58 - 18.27] |
| Hinduism | 37.16 [25.93 - 49.97] | 37.16 [25.93 - 49.97] | 0 |
| Other | 18.42 [10.12 - 31.17] | 18.42 [10.12 - 31.17] | 0 |
| **Functional disability** |  |  |  |
| Person with disability | 24.18 [8.47 - 52.36] | 24.18 [8.47 - 52.36] | 0 |
| Person without disability | 18.22 [10.98 - 28.71] | 18.08 [10.91 - 28.45] | 0.85 [0.20 - 3.52] |
| **Strata** |  |  |  |
| Equateur | 23.83 [16.32 - 33.41] | 23.47 [16.28 - 32.59] | 0.64 [0.19 - 2.09] |
| Kasai | 29.07 [14.65 - 49.47] | 28.99 [14.52 - 49.53] | 3.58 [0.63 - 17.78] |
| Katanga | 23.05 [5.98 - 58.52] | 23.05 [5.98 - 58.52] | 0.29 [0.17 - 0.51] |
| Kivu | 8.57 [2.08 - 29.32] | 8.57 [2.08 - 29.32] | 0.01 [0.00 - 0.04] |
| Leopoldville | 18.49 [15.70 - 21.64] | 18.49 [15.70 - 21.64] | 0 |
| Oriental | 50.43 [37.94 - 62.86] | 49.02 [38.80 - 59.33] | 3.24 [0.60 - 15.55] |
