## Supplementary Material Table 3 for "Prevalence and factors associated with tobacco and nicotine product use among adolescents in the Democratic Republic of the Congo: evidence from a cross-sectional national household survey"

**Supplementary Material Table 3: Prevalence of ever use of smoked tobacco products**

| **Characteristic** | **All smocked tobacco products**  **% (95% CI)** | **Cigarettes** | | | **Shisha**  **% (95% CI)** |
| --- | --- | --- | --- | --- | --- |
|  |  | **Manufactured**  **% (95% CI)** | **Roll-your-own**  **% (95% CI)** | **Any cigarette**  **% (95% CI)** |  |
| **Overall** | 12.28 [6.88 - 20.95] | 9.06 [4.77 - 16.54] | 5.32 [2.60 - 10.57] | 11.30 [6.34 – 19.34] | 2.23 [1.26 - 3.92] |
| **Sex** |  |  |  |  |  |
| Boys | 17.31 [11.33 – 25.55] | 13.96 [8.37 – 22.35] | 7.91 [4.14 – 14.60] | 16.61 [10.72 – 24.83] | 2.91 [1.94 – 4.33] |
| Girls | 6.66 [2.48 – 16.68] | 3.60 [1.27 – 9.74] | 2.42 [0.64 – 8.77] | 5.39 [2.02 – 13.61] | 1.47 [0.43 – 4.94] |
| **Age** |  |  |  |  |  |
| 10 - 12 yrs | 8.84 [5.37 - 14.21] | 5.52 [2.76 - 10.72] | 4.53 [2.07 - 9.65] | 8.61 [5.13 - 14.08] | 0.57 [0.13 - 2.48] |
| 13 -15 yrs | 12.28 [6.35 - 22.40] | 9.34 [4.92 - 17.02] | 4.92 [1.73 - 13.20] | 10.81 [5.51 - 20.11] | 2.79 [1.42 - 5.42] |
| 16 - 17yrs | 20.42 [9.72 - 37.95] | 16.89 [8.12 - 31.87] | 7.96 [4.83 - 12.84] | 18.65 [9.04 - 34.60] | 5.04 [3.25 - 7.72] |
| **Schooling status** |  |  |  |  |  |
| **In-school** | 11.52 [6.31 - 20.12] | 8.18 [4.13 - 15.54] | 4.83 [4.51 - 5.86] | 10.52 [5.80 - 18.35] | 2 [1.12 - 3.54] |
| **Out-of-school** | 17.69 [11.19 - 26.83] | 15.36 [10.00 - 22.87] | 8.77 [11.28 - 17.16] | 16.87 [10.42 - 26.14] | 3.88 [2.29 - 6.48] |
| **Residence** |  |  |  |  |  |
| Rural | 9.89 [5.78 - 16.41] | 7.75 [3.98 - 14.56] | 4.48 [2.43 - 8.11] | 9.59 [5.37 - 16.54] | 1.1 [0.50 - 2.42] |
| Urban | 21.76 [11.82 - 36.60] | 14.24 [7.79 - 24.62] | 8.65 [3.19 - 21.42] | 18.09 [9.93 - 30.67] | 6.7 [2.17 - 18.91] |
| **SES/Wealth index** |  |  |  |  |  |
| 1st quintile: lowest | 14.11 [9.04 - 21.37] | 9.17 [4.80 - 16.82] | 7.48 [3.40 - 15.67] | 13.66 [8.83 - 20.55] | 1.3 [0.29 - 5.63] |
| 2nd quintile: low | 13.41 [9.81 - 18.07] | 11.02 [7.58 - 15.75] | 6.62 [3.81 - 11.28] | 12.79 [8.86 - 18.12] | 3.27 [1.28 - 8.09] |
| 3rd quintile: middle | 11.98 [4.30 - 29.20] | 8.32 [3.20 - 19.95] | 4.7 [1.27 - 15.89] | 10.99 [4.10 - 26.26] | 1.44 [0.26 - 7.60] |
| 4th quintile: high | 11.76 [4.57 - 27.05] | 8.27 [3.00 - 20.79] | 4.61 [1.91 - 10.71] | 10.07 [3.85 - 23.88] | 2.95 [1.08 - 7.79] |
| 5th quintile: highest | 10.07 [5.15 - 18.75] | 8.52 [4.21 - 16.47] | 3.16 [1.32 - 7.37] | 8.93 [4.44 - 17.17] | 2.22 [0.84 - 5.77] |
| **Marital status** |  |  |  |  |  |
| Not in a union | 17.8 [9.46 - 30.96] | 14.49 [7.99 - 24.84] | 7.41 [4.14 - 12.92] | 16.29 [8.92 - 27.88] | 4.04 [2.29 - 7.06] |
| In a union | 19.26 [6.37 - 45.56] | 12.51 [3.05 - 39.42] | 17.09 [5.17 - 43.81] | 19.26 [6.37 - 45.56] |  |
| **Engagement in work** |  |  |  |  |  |
| Employee, n (%) | 28.59 [5.41 - 21.22] | 16.19 [8.82 - 27.85] | 20.46 [14.45 - 28.16] | 26.34 [22.23 - 30.92] | 11.28 [8.11 - 15.50] |
| Self-employed, n (%) | 20.89 [2.54 - 7.43] | 18.43 [5.48 - 46.80] | 7.32 [3.13 - 16.17] | 19.46 [6.16 - 47.08] | 4.65 [1.02 - 18.68] |
| No work, n (%) | 11.04 [68.84 - 73.18] | 8.08 [3.98 - 15.73] | 4.51 [1.59 - 12.13] | 10.11 [4.94 - 19.60] | 1.64 [0.63 - 4.21] |
| **Religion** |  |  |  | Religion |  |
| No religion | 28.3 [16.21 - 44.61] | 25.33 [16.27 - 37.20] | 14.79 [5.54 - 33.94] | 28.3 [16.21 - 44.61] | 1.38 [0.52 - 3.63] |
| Christianity | 11.76 [6.18 - 21.23] | 8.31 [3.96 - 16.61] | 4.92 [2.52 - 9.39] | 10.68 [5.66 - 19.23] | 2.43 [1.34 - 4.35] |
| Islam | 13.48 [3.25 - 41.95] | 12.75 [2.91 - 41.58] | 9.28 [2.56 - 28.51] | 13.22 [3.13 - 41.80] | 0.26 [0.04 - 1.72] |
| Hinduism | 5.49 [0.47 - 41.48] | 5.49 [0.47 - 41.48] |  | 5.49 [0.47 - 41.48] |  |
| Other | 6.16 [2.27 - 15.68] | 5.79 [2.12 - 14.85] | 0.68 [0.22 - 2.07] | 6 [2.21 - 15.25] | 0.28 [0.03 - 2.56] |
| **Functional disability** |  |  |  |  |  |
| Person with disability | 20.53 [6.85 - 47.58] | 14.9 [8.74 - 24.25] | 9.28 [7.66 - 11.21] | 16.46 [7.66 - 31.88] | 12.88 [7.52 - 21.19] |
| Person without disability | 11.82 [6.75 - 19.91] | 8.74 [4.48 - 16.34] | 5.1 [2.29 - 10.98] | 11.02 [6.15 - 18.95] | 1.64 [0.81 - 3.32] |
| **Strata** |  |  |  |  |  |
| Equateur | 9.45 [4.40 - 19.15] | 6.51 [3.13 - 13.05] | 4.29 [1.78 - 9.98] | 9.35 [4.34 - 18.99] | 0.1 [0.01 - 1.20] |
| Kasai | 26.71 [14.44 - 44.04] | 22.62 [13.23 - 35.93] | 12.94 [2.84 - 43.09] | 25.8 [12.89 - 44.98] | 1.82 [0.15 - 19.05] |
| Katanga | 19.78 [5.46 - 51.31] | 18.58 [5.31 - 48.13] | 6.76 [0.91 - 36.51] | 19.53 [5.27 - 51.43] | 1.16 [0.32 - 4.15] |
| Kivu | 7.96 [2.18 - 25.10] | 5.43 [1.09 - 23.02] | 2.81 [1.93 - 4.09] | 6.79 [1.90 - 21.55] | 2.69 [1.17 - 6.04] |
| Leopoldville | 5.34 [3.03 - 9.25] | 3.74 [2.03 - 6.79] | 2.17 [0.92 - 5.03] | 4.65 [2.07 - 10.09] | 1.16 [0.26 - 5.06] |
| Oriental | 23.79 [14.01 - 37.43] | 11.83 [6.40 - 20.82] | 13.62 [3.89 - 38.01] | 21.93 [14.40 - 31.94] | 5.09 [2.87 - 8.87] |
