## Supplementary Material Table 4 for "Prevalence and factors associated with tobacco and nicotine product use among adolescents in the Democratic Republic of the Congo: evidence from a cross-sectional national household survey"

**Supplementary Material Table 4: Prevalence of ever use of smokeless tobacco, heated tobacco and nicotine products**

| **Characteristic** | **Smokeless tobacco products**  **% (95% CI)** | **Heated Tobacco Products**  **% (95% CI)** | **Nicotine pouches**  **% (95% CI)** | **Electronic cigarettes** |
| --- | --- | --- | --- | --- |
| **Overall** | 9.00 [5.60 - 14.14] | 0.17 [0.06 - 0.51] | 0.80 [0.19 - 3.36] | 0.15 [0.02 - 0.98] |
| **Sex** |  |  |  |  |
| Boys | 10.23 [6.82 – 15.06] | 0.28 [0.10 - 0.77] | 1.20 [0.29 - 4.82] | 0.21 [0.03 - 1.67] |
| Girls | 7.62 [4.15 - 13.59] | 0.05 [0.01 - 0.38] | 0.36 [0.07 - 1.82] | 0.36 [0.07 - 1.82] |
| **Age** |  |  |  |  |
| 10 - 12 yrs | 7.2 [4.51 - 11.33] | 0.1 [0.02 - 0.55] | 0.8 [0.12 - 5.29] | 0.02 [0.00 - 0.18] |
| 13 -15 yrs | 9.58 [5.13 - 17.18] | 0.19 [0.07 - 0.51] | 0.59 [0.14 - 2.42] | 0.28 [0.03 - 2.49] |
| 16 - 17yrs | 12.09 [7.61 - 18.68] | 0.29 [0.05 - 1.62] | 1.24 [0.32 - 4.67] | 0.18 [0.04 - 0.77] |
| **Schooling status** |  |  |  |  |
| In-school | 9.31 [5.70 - 14.86] | 0.18 [0.05 - 0.60] | 0.73 [0.51 - 1.04] | 0.17 [0.02 - 1.12] |
| Out-of-school | 6.72 [3.15 - 13.74] | 0.1 [0.02 - 0.47] | 1.37 [0.87 - 3.19] |  |
| **Residence** |  |  |  |  |
| Rural | 7.53 [4.28 - 12.92] | 0.05 [0.01 - 0.22] | 0.29 [0.04 - 2.26] |  |
| Urban | 14.81 [8.64 - 24.22] | 0.66 [0.18 - 2.37] | 2.84 [0.78 - 9.80] | 0.73 [0.12 - 4.18] |
| **Wealth quintile** |  |  |  |  |
| 1st quintile: lowest | 9.36 [3.73 - 21.58] |  | 0.81 [0.11 - 5.78] | 0.06 [0.01 - 0.56] |
| 2nd quintile: low | 11.7 [4.91 - 25.40] |  | 0.74 [0.12 - 4.53] | 0.03 [0.00 - 0.26] |
| 3rd quintile: middle | 9.34 [4.37 - 18.86] | 0.2 [0.06 - 0.65] | 1.06 [0.20 - 5.32] | 0.11 [0.02 - 0.68] |
| 4th quintile: high | 6.5 [4.43 - 9.44] | 0.41 [0.07 - 2.36] | 0.9 [0.14 - 5.47] | 0.53 [0.06 - 4.58] |
| 5th quintile: highest | 7.94 [5.16 - 12.03] | 0.23 [0.04 - 1.16] | 0.48 [0.09 - 2.59] |  |
| **Marital status** |  |  |  |  |
| Not in a union | 11.92 [7.29 - 18.89] | 0.27 [0.06 - 1.27] | 1.08 [0.25 - 4.48] | 0.35 [0.05 - 2.43] |
| In a union | 18.68 [3.28 - 60.86] | 1.14 [0.11 - 10.99] | 4.84 [0.99 - 20.65] |  |
| **Engagement in work** |  |  |  |  |
| Employee, n (%) | 4.92 [0.70 - 27.58] | 1.77 [0.19 - 14.35] | 1.02 [0.09 - 10.48] |  |
| Self-employed, n (%) | 9.07 [4.05 - 19.07] | 0.1 [0.02 - 0.69] | 1.33 [0.27 - 6.32] | 0.26 [0.04 - 1.78] |
| No work, n (%) | 9.51 [5.53 - 15.88] | 0.11 [0.03 - 0.42] | 0.77 [0.16 - 3.61] | 0.15 [0.02 - 1.04] |
| **Religion** |  |  |  |  |
| No religion | 18.63 [9.47 - 33.36] | 0.19 [0.03 - 1.25] | 5.13 [1.00 - 22.50] |  |
| Christianity | 8.09 [5.02 - 12.78] | 0.11 [0.02 - 0.52] | 0.54 [0.13 - 2.26] | 0.16 [0.02 - 1.11] |
| Islam | 12.67 [4.29 - 31.97] | 1.81 [0.22 - 13.32] | 3.49 [0.58 - 18.27] |  |
| Hinduism | 31.67 [12.72 - 59.58] |  |  |  |
| Other | 17.34 [9.10 - 30.52] | 0.27 [0.04 - 1.98] |  |  |
| **Functional disability** |  |  |  |  |
| Person with disability | 4.17 [1.04 - 15.23] |  |  |  |
| Person without disability | 9.26 [5.81 - 14.45] | 0.18 [0.06 - 0.53] | 0.85 [0.20 - 3.52] | 0.15 [0.02 - 1.04] |
| **Strata** |  |  |  |  |
| Equateur | 18.53 [12.90 - 25.89] | 0.12 [0.01 - 1.37] | 0.64 [0.19 - 2.09] | 0.46 [0.06 - 3.18] |
| Kasai | 11.59 [2.76 - 37.68] | 0.5 [0.14 - 1.77] | 3.58 [0.63 - 17.78] |  |
| Katanga | 5.1 [0.88 - 24.58] | 0.38 [0.06 - 2.51] | 0.29 [0.17 - 0.51] |  |
| Kivu | 0.75 [0.04 - 11.59] |  | 0.01 [0.00 - 0.04] | 0.01 [0.00 - 0.04] |
| Leopoldville | 15.11 [13.30 - 17.11] | 0.01 [0.00 - 0.14] |  |  |
| Oriental | 35.38 [29.26 - 42.01] | 0.8 [0.15 - 4.08] | 3.24 [0.60 - 15.55] | 1.57 [0.30 - 7.86] |
