## Supplementary Material Table 5 for "Prevalence and factors associated with tobacco and nicotine product use among adolescents in the Democratic Republic of the Congo: evidence from a cross-sectional national household survey"

**Supplementary Material Table 5: Prevalence of current use of heated tobacco products**

|  | **n** | **% [95% CI]** |
| --- | --- | --- |
| **overall** | 09 | 0.11 [0.11-0.11] |
| **Sex** | | |
| Boys | 09 | 0.11 [0.11-0.11] |
| Girls |  |  |
| **Age** |  |  |
| 10 - 12 yrs | 5 | 0.10 [0.10 - 0.10] |
| 13 -15 yrs | 1 | 0.03 [0.03 - 0.03] |
| 16 - 17yrs | 3 | 0.29 [0.28 - 0.29] |
| **Schooling status** |  |  |
| In-school | 8 | 0.12 [0.12 - 0.12] |
| Out-of-school | 1 | 0.04 [0.03 - 0.04] |
| **Residence (rural/urban)** |  |  |
| Rural | 5 | 0.04 [0.04 - 0.04] |
| Urban | 4 | 0.38 [0.38 - 0.39] |
| **SES/Wealth index** |  |  |
| 1st quintile: lowest | 0 | 0.0 |
| 2nd quintile: low | 0 | 0.0 |
| 3rd quintile: middle | 1 | 0.03 [0.03 - 0.03] |
| 4th quintile: high | 5 | 0.30 [0.30 - 0.31] |
| 5th quintile: highest | 3 | 0.22 [0.21 - 0.22] |
| **Marital status** |  |  |
| Not in a union | 3 | 0.17 [0.17 - 0.18] |
| In a union | 1 | 1.14 [1.08 - 1.21] |
| Separated | 0 | 0.0 |
| Partner deceased | 0 | 0.0 |
| **Engagement in work** |  |  |
| Employee, n (%) | 2 | 1.26 [1.24 - 1.28] |
| Self-employed, n (%) | 0 | 0.0 |
| No work, n (%) | 7 | 0.07 [0.07 - 0.07] |
| **Religion** |  |  |
| No religion | 2 | 0.19 [0.18 - 0.20] |
| Christianity | 5 | 0.08 [0.08 - 0.08] |
| Islam | 2 | 1.04 [1.01 - 1.06] |
| Hinduism | 0 | 0.0 |
| Other | 0 | 0.0 |
| **Functional disability** |  |  |
| Person with disability | 0 | 0.0 |
| Person without disability | 9 | 0.11 [0.11 - 0.11] |
| **Strata** |  |  |
| Equateur | 1 | 0.12 [0.11 - 0.13] |
| Kasai | 3 | 0.28 [0.27 - 0.28] |
| Katanga | 4 | 0.28 [0.27 - 0.28] |
| Kivu | 0 | 0.0 |
| Leopoldville | 0 | 0.0 |
| Oriental | 1 | 0.53 [0.52 - 0.54] |
