## Supplementary Material Table 6 for "Prevalence and factors associated with tobacco and nicotine product use among adolescents in the Democratic Republic of the Congo: evidence from a cross-sectional national household survey"

**Supplementary Material Table 6: Prevalence of current use of smokeless tobacco products**

| **Characteristic** | **Prevalence**  **% (95% CI)** |
| --- | --- |
| **Overall** | 5.86 [3.42 - 9.87] |
| **Sex** |  |
| Boys | 6.71 [3.81 – 11.56] |
| Girls | 4.91 [2.78 – 8.54] |
| **Age** |  |
| 10 - 12 yrs | 5.31 [2.86 - 9.67] |
| 13 -15 yrs | 5.63 [2.89 - 10.69] |
| 16 - 17yrs | 7.61 [4.66 - 12.21] |
| **Schooling status** |  |
| In-school | 5.97 [3.53 - 9.92] |
| Out-of-school | 5.1 [1.93 - 12.79] |
| **Residence** |  |
| Rural | 4.99 [2.88 - 8.51] |
| Urban | 9.33 [4.17 - 19.57] |
| **SES/Wealth index** |  |
| 1st quintile: lowest | 6.4 [2.36 - 16.23] |
| 2nd quintile: low | 7.9 [3.47 - 16.98] |
| 3rd quintile: middle | 6.33 [2.84 - 13.51] |
| 4th quintile: high | 3.98 [2.24 - 6.98] |
| 5th quintile: highest | 4.56 [2.28 - 8.93] |
| **Marital status** |  |
| Not in a union | 7.32 [4.44 - 11.82] |
| In a union | 7.56 [2.20 - 22.89] |
| **Engagement in work** |  |
| Employee, n (%) | 3.7 [0.55 - 21.00] |
| Self-employed, n (%) | 8.07 [3.32 - 18.34] |
| No work, n (%) | 6.05 [3.38 - 10.61] |
| **Religion** |  |
| No religion | 13.21 [4.28 - 34.10] |
| Christianity | 5.18 [3.24 - 8.19] |
| Islam | 11.06 [2.96 - 33.62] |
| Hinduism | 31.67 [12.72 - 59.58] |
| Other | 7.9 [2.47 - 22.48] |
| **Functional disability** |  |
| Person with disability | 0.18 [0.02 - 1.45] |
| Person without disability | 6.17 [3.65 - 10.27] |
| **Strata** |  |
| Equateur | 10.24 [6.04 - 16.85] |
| Kasai | 10.09 [2.26 - 35.24] |
| Katanga | 4.31 [0.75 - 21.11] |
| Kivu | 0.18 [0.01 - 2.92] |
| Leopoldville | 8.02 [5.44 - 11.66] |
| Oriental | 25.03 [20.67 - 29.96] |
