## Supplementary Material Table 7 for "Prevalence and factors associated with tobacco and nicotine product use among adolescents in the Democratic Republic of the Congo: evidence from a cross-sectional national household survey"

**Supplementary Material Table 7: Prevalence of current use of nicotine products**

| **Characteristic** | **All nicotine products**  **(pouches + e-cigarettes)**  **% (95% CI)** | **Nicotine pouches**  **% (95% CI)** | **Electronic cigarettes**  **% (95% CI)** |
| --- | --- | --- | --- |
| **Overall** | 0.60 [0.10 - 3.40] | 0.56 [0.09 - 3.55] | 0.04 [0.01 - 0.16] |
| **Sex** |  |  |  |
| Boys | 0.84 [0.15 - 4.64] | 0.83 [0.14 - 4.67] | 0.01 [0.00 - 0.05] |
| Girls | 0.33 [0.06 - 1.90] | 0.26 [0.03 - 2.31] | 0.07 [0.02 - 0.37] |
| **Age** |  |  |  |
| 10 - 12 yrs | 0.72 [0.09 - 5.71] | 0.7 [0.08 - 5.88] | 0.02 [0.00 - 0.18] |
| 13 -15 yrs | 0.28 [0.04 - 1.85] | 0.28 [0.04 - 1.85] |  |
| 16 - 17yrs | 0.93 [0.23 - 3.70] | 0.77 [0.17 - 3.33] | 0.16 [0.03 - 0.76] |
| **Schooling status** |  |  |  |
| In-school | 0.51 [0.09 - 2.71] | 0.46 [0.07 - 2.84] | 0.04 [0.01 - 0.19] |
| Out-of-school | 1.26 [0.17 - 8.65] | 1.26 [0.17 - 8.65] |  |
| **Residence** |  |  |  |
| Rural | 0.28 [0.03 - 2.39] | 0.28 [0.03 - 2.39] |  |
| Urban | 1.84 [0.39 - 8.30] | 1.65 [0.30 - 8.60] | 0.19 [0.05 - 0.75] |
| **Wealth quintile** |  |  |  |
| 1st quintile: lowest | 0.73 [0.08 - 6.41] | 0.67 [0.06 - 6.91] | 0.06 [0.01 - 0.56] |
| 2nd quintile: low | 0.66 [0.08 - 4.98] | 0.64 [0.08 - 5.08] | 0.01 [0.00 - 0.14] |
| 3rd quintile: middle | 0.95 [0.15 - 5.74] | 0.84 [0.10 - 6.41] | 0.11 [0.02 - 0.68] |
| 4th quintile: high | 0.15 [0.02 - 0.96] | 0.15 [0.02 - 0.96] |  |
| 5th quintile: highest | 0.46 [0.08 - 2.63] | 0.46 [0.08 - 2.63] |  |
| **Marital status** |  |  |  |
| Not in a union | 0.65 [0.16 - 2.64] | 0.56 [0.13 - 2.49] | 0.09 [0.02 - 0.42] |
| In a union | 4.84 [0.99 - 20.65] | 4.84 [0.99 - 20.65] |  |
| **Engagement in work** |  |  |  |
| Employee, n (%) |  |  |  |
| Self-employed, n (%) | 1.3 [0.25 - 6.40] | 1.07 [0.23 - 4.90] | 0.23 [0.03 - 1.99] |
| No work, n (%) | 0.57 [0.07 - 4.25] | 0.55 [0.07 - 4.43] | 0.02 [0.00 - 0.13] |
| **Religion** |  |  |  |
| No religion | 5.13 [1.00 - 22.50] | 5.13 [1.00 - 22.50] |  |
| Christianity | 0.33 [0.07 - 1.50] | 0.28 [0.05 - 1.52] | 0.04 [0.01 - 0.18] |
| Islam | 3.11 [0.37 - 21.55] | 3.11 [0.37 - 21.55] |  |
| Hinduism |  |  |  |
| Other |  |  |  |
| **Functional disability** |  |  |  |
| Person with disability |  |  |  |
| Person without disability | 0.63 [0.11 - 3.57] | 0.59 [0.09 - 3.72] | 0.04 [0.01 - 0.17] |
| **Strata** |  |  |  |
| Equateur | 0.56 [0.18 - 1.74] | 0.18 [0.02 - 2.05] | 0.38 [0.05 - 2.67] |
| Kasai | 3.42 [0.54 - 18.85] | 3.42 [0.54 - 18.85] |  |
| Katanga | 0.09 [0.01 - 0.90] | 0.09 [0.01 - 0.90] |  |
| Kivu | 0.01 [0.00 - 0.04] |  | 0.01 [0.00 - 0.04] |
| Leopoldville |  |  |  |
| Oriental | 1.18 [0.22 - 5.95] | 0.91 [0.17 - 4.64] | 0.27 [0.05 - 1.38] |
